# Analysis of Delirium in Intensive Care Patients using Bayesian Networks

**DOI:** 10.1101/2025.09.23.25336328

**Authors:** Jasmin Bachmann, Marcos Delgado, Reto A. Schuepbach, Jan Bartussek, Georg R. Spinner

**Author notes:** **Corresponding author:** Georg Spinner.

## Abstract

Delirium is a multifactorial and complex syndrome commonly observed in intensive care unit (ICU) patients, significantly affecting outcomes, mortality, and healthcare costs. Despite numerous potential risk factors, its exact pathophysiology remains unclear. This study explores the use of Additive Bayesian Networks (ABNs) to analyse ICU patient data and identify delirium risk factors. Three cohorts with delirium-related symptoms were examined: general delirium, delirium tremens (DT), and hepatic encephalopathy (HE). Delirium was measured by the Intensive Care Delirium Screening Checklist (ICDSC) whereas the other cohort depend on the coded diagnosis. The analysis shows important connections, such as the two-way link between antipsychotic medication and delirium and the connections of ventilation and age to delirium. Some of the results are in line with research like the link between high levels of ammonia and HE and the strong link between infection, painkillers, and pain that remains the same across all three groups. However, novel insights, such as the reduced likelihood of delirium (ICDSC ≥4) with age and ventilation, challenge conventional understanding. This study shows how ABNs can be used to find complicated dependencies in the ICU. Future work should focus on addressing potential overfitting or unmodeled interactions in the dataset and exploring specific delirium subtypes, such as hypoactive and hyperactive delirium.

## Introduction

### Delirium

Delirium is an acute, fluctuating, and mostly reversible syndrome characterised by a combined disturbance of attention and orientation, along with the impairment of another cognitive function (Hermes et al., 2022). It is one of the most common postoperative complications in older adults, and more than half of all patients in intensive care units (ICUs) develop delirium during their stay (Mart et al., 2021). Overall, 30–80% of ICU patients and 5–52% of surgical patients, depending on the procedure, experience delirium. In high-risk surgeries, such as trauma and heart operations, the prevalence is 36– 40%, whereas in low-risk procedures, including elective arthroplasty, 5–10% of adults develop postoperative delirium (Swarbrick & Partridge, 2022).

This syndrome is associated with increased mortality, prolonged hospitalisation, and poorer outcomes. Approximately 25% of patients retain cognitive dysfunction after delirium (Zoremba & Coburn, 2019). Moreover, the management of patients with delirium adds to the burden on nursing and clinical staff in the already fast-paced ICU environment (Hermes et al., 2022). Beyond these immediate challenges, there are also long-term consequences: delirium has been linked to prolonged cognitive impairment, which can significantly affect cardiac rehabilitation efforts (Sato et al., 2017).

Delirium can be categorised into hyperactive, hypoactive, and mixed delirium. Hyperactive delirium is characterised by heightened alertness and uncooperative behaviour, whereas hypoactive delirium, which is more prevalent than the hyperactive form, is marked by excessive sleep, disorganised thinking, and inattention. Mixed delirium, the most commonly observed subtype, involves fluctuating patterns between hyperactivity and hypoactivity.

Hyperactive delirium patients often pose a risk of harm to themselves and others due to their combative behaviour, potentially delaying optimal invasive care. Conversely, hypoactive delirium patients may develop complications related to immobility, such as infections and pressure sores (Van Lieshout et al., 2022), (Sato et al., 2017, p. 557).

Delirium is difficult to diagnose and often overlooked (Tran et al., 2021). Validated screening tools for delirium in the ICU include the Confusion Assessment Method for the ICU (CAM-ICU) and the Intensive Care Delirium Screening Checklist (ICDSC). The CAM-ICU is considered the most reliable for ICU patients, with a sensitivity of 0.79 and a specificity of 0.97. The ICDSC has a high sensitivity of 0.99 and a specificity of 0.64. It can be completed within minutes, is suitable for ventilated patients, and can detect subsyndromal delirium (Luetz et al., 2010), (‘Screening for Delirium with the Intensive Care Delirium Screening Checklist (ICDSC)’, 2018). The ICDSC serves as a screening tool to identify delirium in patients based on the score they achieve. The items include the assessment of consciousness, orientation, hallucinations or delusions, psychomotor activity, inappropriate speech or mood, attentiveness, sleep-wake cycle disturbances, and fluctuation in symptomatology. The maximum score is 8, and scores of 4 or higher indicate the presence of delirium (‘Screening for Delirium with the Intensive Care Delirium Screening Checklist (ICDSC)’, 2018). However, delirium screening, particularly for the often under-recognised hypoactive delirium subtype, is rarely performed consistently in European hospitals and ICUs. Studies indicate that only 27% of ICU patients are screened with a validated method. Considering that hypoactive delirium has a higher mortality rate than the hyperactive form (33% vs. 15%), the S3 guideline recommends screening ICU patients at least every eight hours (Zoremba & Coburn, 2019) (S3-Leitlinie Analgesie, Sedierung Und Delirmanagement in Der Intensivmedizin (DAS-Leitlinie), 2021).

The causes of delirium are diverse and complex, resulting primarily from direct damage to the central nervous system and adverse stress reactions. Direct brain damage can arise from energy deficiencies (e. g. hypoxia and hypoglycaemia), metabolic disorders (e.g. hyponatraemia and hypercalcaemia), neoplasms, and drug toxicity (Swarbrick & Partridge, 2022). To date, no validated or clinically established biomarkers for delirium exist, making it difficult to diagnose, and it is often overlooked. Among patients over 65 years old, given the more frequent background of dementia or depression, delirium is not identified in almost 30% of cases (Hermes et al., 2022). Given that delirium in ICUs significantly worsens prognosis and increases mortality, research into this disorder is critical (Tran et al., 2021).

As many delirium cases are preventable, it is essential to identify patients at risk and implement targeted preventive measures. These individuals must be closely monitored, and any modifiable predisposing and triggering factors need to be minimised (Mann, 2018). The early identification of patients at increased risk of delirium creates an opportunity to adjust these factors and support effective shared decision-making processes.

Different standard operation models recommend the following steps for preventing and treating delirium. In the ICU setting, adherence to interventions, such as early breathing trials, the restrictive use of analgesia and sedation, regular delirium assessments, early mobilisation, and active family engagement has been shown to reduce the occurrence of delirium. A key strategy for all ICU patients involves employing non-pharmacological measures. This includes implementing activating interventions during the daytime and sleep-promoting interventions at night. Pharmacological delirium prevention should generally be avoided, as studies have shown that agents like haloperidol do not lower delirium incidence or duration, nor do they increase delirium- or coma-free days. Additionally, inappropriate or excessive sedation, which hinders patient communication and contact, should be prevented. Non-pharmacological approaches, namely nursing, nursing-therapeutic, and therapeutic interventions, should be integrated and emphasised. Overall, combining multiple non-pharmacological strategies is more effective than single measures in reducing delirium, ventilation time, length of stay, and possibly mortality (S3-Leitlinie Analgesie, Sedierung Und Delirmanagement in Der Intensivmedizin (DAS-Leitlinie), 2021), (Hermes et al., 2022), (Kukolja & Kuhn, 2021).

### Aim

In recent years, several studies have focused on the prediction of delirium using other machine learning methods. A prediction model used three machine learning algorithms: logistic regression, random forest, and gradient boosting (Gong et al., 2023). Another study employed a large language model to predict delirium, utilizing EHR data available within the first 24 hours of ICU admission (Contreras et al., 2024). However, new studies analysing delirium using a Bayesian network (BN) to reveal the connection between the influencing factors have not been published.

Therefore, this study’s aims to identify the relationships between predisposing and precipitating risk factors for delirium. Because some factors may not directly cause delirium but may instead affect other factors that lead to delirium, the hypothesis is that connections between multiple factors exist that have not been seen before. To benchmark the model, the well-known disease hepatic encephalopathy is analysed and then compared with delirium and delirium tremens. By comparing the resulting network structures, the aim is to determine whether similar underlying relationships between these different but symptomatically overlapping diseases are evident.

## Methods

In order to find information on the links between different risk factors for delirium, it was first necessary to research the possible risk factors. The most common risk factors, which were also available in a structured form, were selected. Then the data was extracted from the database and processed to obtain the appropriate features. Finally, the Additive Bayesian Network (ABN) analysis was performed in R.

### Data

The dataset consisted of retrospective patient data from the ICU of the university hospital in Zurich.

They were extracted from the clinical information system of the ICU from a research SQL database and then processed in a research server in R-Studio 2024 with R Version 4.4.1.

### Risk Factors

Because many delirium cases are preventable, implementing effective preventive measures is critical. A key step in developing these strategies is identifying vulnerable patients by assessing both their predisposing and foreseeable triggering factors (Mann, 2018).

Predisposing factors, including advanced age (>65 years), male gender, dementia and other cognitive disorders, depression, prior delirium, sensory impairments, multiple medications, multiple comorbidities, and poor general and nutritional status, establish a baseline risk (Mann, 2018), (Qureshi & Arthur, 2023), (Ali & Cascella, 2024), (AMBOSS GmbH, n.d.). Triggers such as medication use, poorly treated pain, infections, prolonged mechanical ventilation, hypoxia, and various metabolic imbalances (e.g. hypo-/hyperglycaemia) can precipitate delirium onset (Qureshi & Arthur, 2023), (AMBOSS GmbH, n.d.).

Several observational studies have corroborated these risk factors. According to Arizumi et al., 2021 the most important risk factors for postoperative delirium are psychiatric disorders, benzodiazepine use, age over 70 years, hearing loss, and ICU admission. Additionally, the administration of opioids is part of intra- and post-operative care, whereas opioids can increase the risk of delirium; likewise, pain managed by opioids is also a risk factor (Duprey et al., 2021). One observational study revealed a significant association of delirium with nasogastric intubation, urinary catheterisation, tracheal intubation, and invasive mechanical ventilation (Tran et al., 2021).

Given the close relationship between delirium, agitation, and pain, assessing these factors was deemed necessary (Zoremba & Coburn, 2019, p. 103). Three pain scores are being measured at the University Hospital of Zurich: the Verbal Rating Scale (VRS), the Numeric Rating Scale (NRS), and the Zurich Observation Pain Assessment (ZOPA). Notably, these scores are diverse: VRS (0–5) rates pain based on descriptive adjectives, NRS (0–10) assigns it a numerical value, and ZOPA captures behavioural signs of pain without quantifying the intensity. The VRS and NRS values are assessed by the patient, whereas ZOPA is analysed by medical personnel, and it records pain in four behavioural categories: vocalisations, facial expressions, body language, and physiological indicators (Thomm, 2016),(ZOPA© – Das Zurich Observation Pain Assessment, n.d.).

Simplified Acute Physiology Score (SAPS) II, which predicts hospital mortality, was used to supplement previous comorbidities as risk factors. It uses 17 variables: 12 are physiological variables, and the others are age, type of admission (scheduled surgical, unscheduled surgical, or medical), and three underlying diseases with AIDS, metastatic cancer, and hematologic malignancy. With the physiologic variables the worst values of the first 24 hours in the ICU are used (Afessa et al., 2007).

Although multiple potential risk factors were identified in the literature, only some variables were ultimately selected for the following analyses. This choice was guided by the structured availability of data, data quality, and the need to avoid highly correlated variables.

### Preprocessing

In BNs, the computing time required for calculating feature probabilities or finding optimal network structures depends heavily on certain parameters. Particularly, the time required increases significantly with the maximum parent degree. In addition, the memory requirement of the approach involves using scales exponentially, the number of which practically limits the method to a maximum of approximately 25 variables (Koivisto & Sood, 2004).

Therefore, data preprocessing is crucial to minimise the number of features. Because this study focuses on identifying factors that influence delirium, data from the 24 hours preceding the onset of delirium were examined. Different methods were tested to determine the onset and define a common timepoint (see Appendix 5). Specifically, the first instance of delirium corresponded to an ICDSC score exceeding 3 on two consecutive measurements, with no Richmond Agitation-Sedation Scale (RASS) below −3 within 10 min of the ICDSC measurement. According to standard operating procedures, the ICDSC should not be measured when RASS is below −3, as delirium is then not assessable (S3-Leitlinie Analgesie, Sedierung Und Delirmanagement in Der Intensivmedizin (DAS-Leitlinie), 2021). The first measurement exceeding an ICDSC score of 3 was then designated as the timepoint of delirium. To approximate a comparable timepoint for non-delirious patients, their mean ICDSC measurement was used if available or else, the last recorded measurement was used.

According to an observational study, the first symptoms of delirium in patients occurred 17.9 ± 34.1 h after hospitalisation and lasted 4.9±4.9 days (Tran et al., 2021). In this study, the 8^th^ ICDSC measurement (see Table 1) was selected as the timepoint— corresponding to maximum 64 hours, since University Hospital Zurich (USZ) conducts at least one ICDSC measurement every 8-hour shift. This indicates that delirium symptoms appear on average after 64 hours since the beginning of the ICU stay.

**Table 1:**
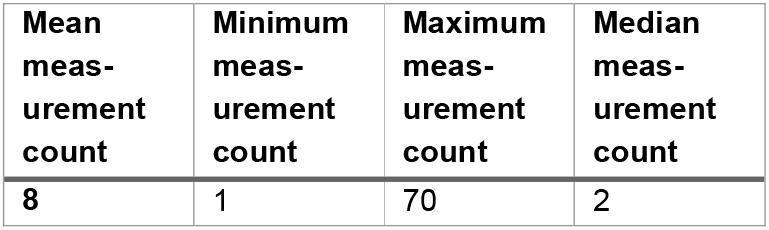
Statistics of the no. of consecutive ICDSC assessments until ICDSC > 3.

| Mean measurement count | Minimum measurement count | Maximum measurement count | Median measurement count |
| --- | --- | --- | --- |
| 8 | 1 | 70 | 2 |

### Feature Development

All laboratory, Arterial Blood Gas (ABG) test, pain, and scoring data from the 24 hours before this threshold was reached were included. Because International Statistical Classification of Diseases and Related Health Problems (ICD) coded data are not timestamped, they could not be filtered by time. SAPS measurements, which are typically recorded at admission, were included up to 14 days before the threshold to ensure sufficient data coverage while still capturing relatively current assessments.

First, the data were extracted from the clinical information system using SQL, where an initial preprocessing step was performed, including combining ICD codes, identifying hyperoxia and hypoxia, and filtering the 24 h timeframe. Subsequent feature creation and data merging were conducted in RStudio. During these stages, generative AI tools were employed for code creation and optimisation, specifically OpenAI’s GPT-4 model (OpenAI, 2023). After finalising these processes, the resulting dataset was then split into three subsets for further analysis.

Medications associated with delirium and commonly used in this ICU were grouped into analgesics, anaesthetics, and antipsychotics. If any medication within these categories was administered 24 hours prior to the defined threshold, it was recorded as given for that category. A comprehensive list of these medications can be found in the Appendix, Chapter 3.

For the laboratory and oxidation variables, the following thresholds (see Table 2) for the different value sets were taken, according to internal thresholds and information from the online medical compendium Amboss (AMBOSS GmbH, 2024b), (AMBOSS GmbH, 2024a), (AMBOSS GmbH, 2024d) and (PADILLA OSVALDO & ABADIE JUDE, 2021).

**Table 2:** Thresholds for laboratory, ABG and oxidation variables.

| Feature | Criteria |
| --- | --- |
| **Hypoxia** | normal >10 partial pressure of oxygen (PaO<sub>2</sub>)<br>low 9–10 PaO<sub>2</sub><br>moderate 8–6.5 PaO<sub>2</sub><br>severe <= 6.6 PaO<sub>2</sub> |
| **Hyperoxia** | normal > 16 PaO<sub>2</sub><br>low 16–19 PaO<sub>2</sub><br>moderate 20–29 PaO<sub>2</sub><br>severe >= 30 PaO<sub>2</sub> |
| **Hypocalcaemia** | ionised calcium <1,15 µmol /L |
| **Hypercalcaemia** | ionised calcium >1,35 µmol /L |
| **Hypokalaemia** | Serum potassium <3,6 µmol /L |
| **Hyperkalaemia** | Serum potassium >5,2 µmol /L |
| **Hyponatremia** | Serum concentration <135 µmol /L |
| **Hypernatremia** | Serum concentration >145 µmol /L |
| **Hyperalbuminemia** | >49 g/L |
| **Hypoalbuminemia** | <40 g/L |
| **High Ammonia** | >47 µmol/l |
| **Low Ammonia** | < 23 µmol/l |

**Table 3:** ICD Codes used for feature design.

| Feature | Criteria |
| --- | --- |
| **ICD Delirium** | ICD Code F05 and subgroups |
| **ICD Delirium Tremens** | ICD Code F10.4 and subgroups |
| **ICD Hepatic encephalopathy** | ICD Code K72.7 and subgroups |
| **ICD Dementia** | ICD Code F00, F01, F02, F03, F06.7 and |

Table 3: ICD Codes used for feature design
|  | subgroups |
| --- | --- |
| **ICD Thyroid** | ICD Code E00, E01, E02, E03, E04, E05, E06, E07 and subgroups |
| **ICD Malnutrition** | ICD Code E40, E41, E42, E43, E44, E45, E46 and subgroups |
| **ICD Diabetes** | ICD Code E10, E11, E12, E13, E14 and subgroups |

Patients with available fraction of inspired oxygen (FiO2) measurements during oxygenation were classified as ventilated, indicating they received mechanical ventilation. As procalcitonin (PCT) measurements were not available for all the patients, this feature was categorised as ‘measured’ and ‘not measured’. PCT is often used as a biomarker specific for bacterial infections in a variety of clinical settings including intensive care (Samsudin & Vasikaran, 2017). The ICDSC score was discretised in the category of no delirium with scores between 0 and 2, possible delirium with a score of 3 and delirium at standard threshold with a score of 4 and higher. Then the standard threshold at the moment is ≥ 4 for delirium, but a study shows that decreasing the ICDSC threshold for delirium to ≥3 increases the accuracy in detecting delirium at the cost of over-identification (‘Screening for Delirium with the Intensive Care Delirium Screening Checklist (ICDSC)’, 2018).

The diagnosis features were extracted using the corresponding ICD codes for these cases and patients:

Pain was assessed using all three standard measurements at USZ, as different personnel and units use diverse methods. Pain was recorded if NRS exceeded 3, VRS indicated severe or strongest pain, or ZOPA results were positive.

The minimum data set of the SGI (MDSI) defines and records key facts and figures for all ICUs in Switzerland. One of these records is the diagnosis/problem at admission (Schweizerische Gesellschaft für Intensivmedizin, n.d.). This feature is used in this analysis to determine the reason why the patient is in the intensive care unit, for example, because of cardiovascular problems. SAPS and age were not further pre-processed, as the abn() package can handle such continuous data, especially if their empirical distributions (see Figure 1) can be approximated by Gaussian distributions.

**Figure 1:**
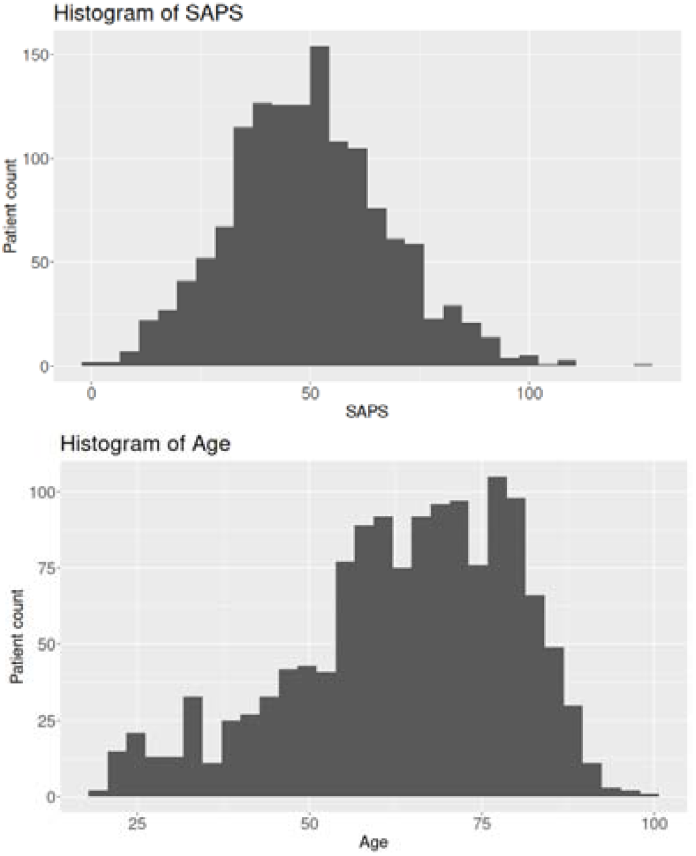
Empirical distributions of SAPS and age

### Cohorts with delirious symptoms

Delirium due to hepatic encephalopathy (HE) and delirium tremens (DT) can manifest with delirious symptoms, potentially showing in elevated scores in ICDSC (Kaye et al., 2024). In this study, three subsets of a delirium cohort were compared: one comprising cases of delirium, another of DT, and a third of HE. While delirium and DT have a comparable range of variables, the HE subgroup contains fewer data points because the ammonia concentration is required there, which is measured in only a fraction of patients. The primary objective is to apply an ABN analysis to each subset to identify influential factors associated with these conditions.

DT results from a complex interaction of neurochemical imbalances triggered by sudden alcohol withdrawal, especially altered GABA and glutamate signalling. This shift, characterised by reduced GABAergic inhibition and increased glutamatergic excitation, leads to neuronal hyperexcitability, excitotoxicity, and typical symptoms of DT, such as confusion, hallucinations, and tremors (Kaye et al., 2024).

HE occurs in individuals with chronic liver disease and can be triggered by various stressors, such as gastrointestinal bleeding, infections, or improper medication use. Patients typically present with confusion, disorientation, sleepiness, personality changes, and mood swings. Contributing factors include infections, prescribed medications, gastrointestinal bleeding, dehydration, electrolyte imbalances, and the use of certain substances like alcohol, sedatives, painkillers, or diuretics (Danielle Tholey, 2023).

While delirium and encephalopathies share some overlapping symptoms, such as cognitive disturbances, they differ significantly in their underlying causes, treatment, and prognosis. Encephalopathies can mimic delirium in attention and memory disruptions but may also present unique features like myoclonus or nystagmus (Piel et al., 2022).

Delirium is not a disease, but a clinical condition characterised by a combination of features, and it is sometimes an expression of acute encephalopathy. The latter is not a clinical syndrome but is understood as a rapidly evolving, diffuse pathobiological process that may manifest as delirium or, in cases of severely impaired consciousness, coma. Against this background, it is important to regularly check whether delirium and/or acute encephalopathy are present (Hermes et al., 2022).

To compare delirium, delirium tremens and hepatic encephalopathy, the original cohort was divided into three groups, excluding the positive diagnoses of the other groups in each group, see Figure 2.

**Figure 2:**
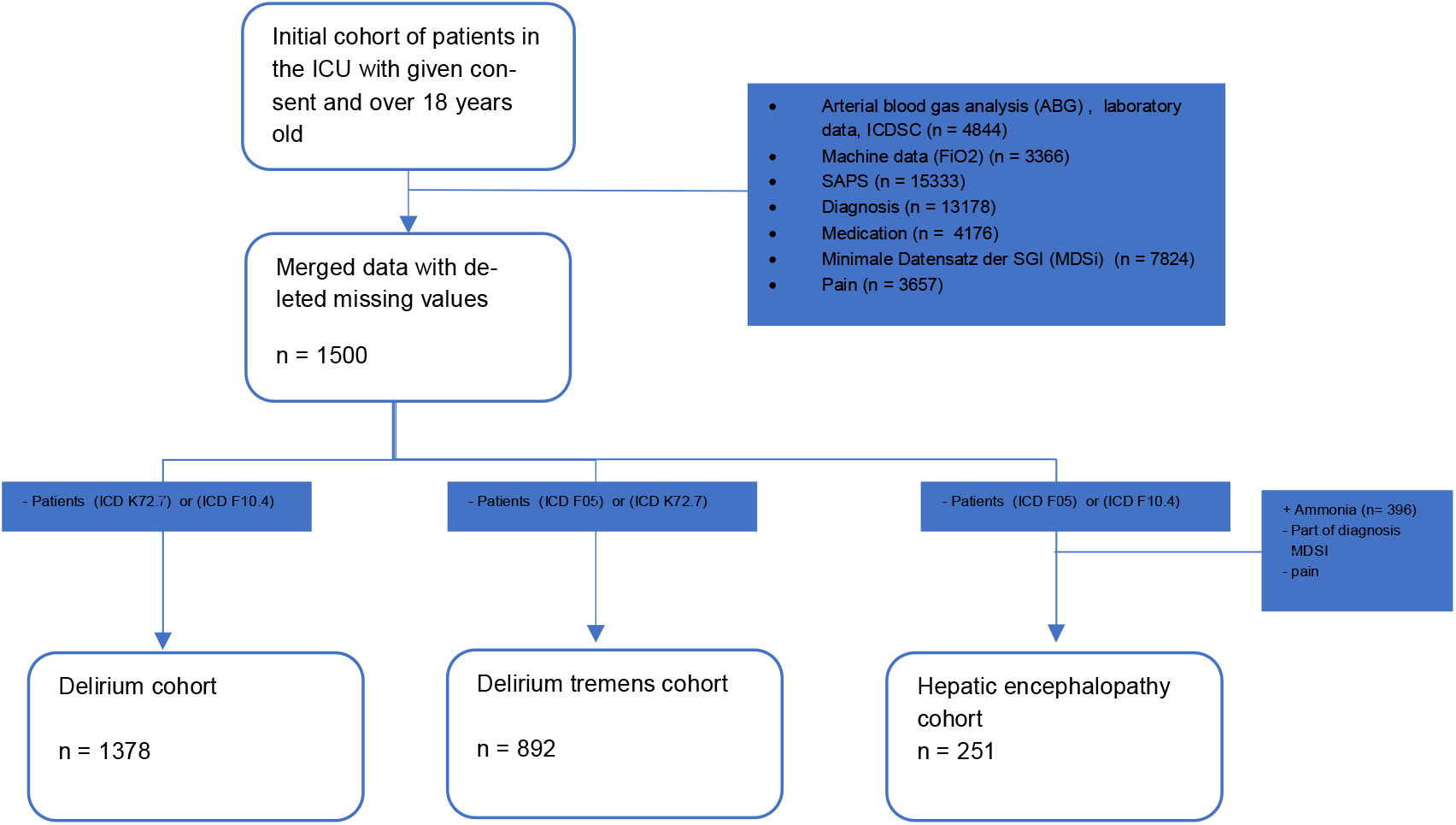
Patient cohorts with exclusion criteria and defining factors

### Bayesian Network

BN modelling has been applied to medical conditions with high degrees of uncertainty in various domains, such as breast cancer, COVID-19, intracranial aneurysm, and multiple sclerosis (Jiang et al., 2019), (Fenton et al., 2020), (Hartmann et al., 2022), (Delucchi et al., 2022). BN offers a probabilistic approach in settings characterised by uncertainty and a need for explainability and trustworthiness (Madsen & Weidl, 2024). One other of the main advantages of BN is the input of domain knowledge through whitelisting and blacklisting, as typically, some connections are already known (Furrer, R. et al., 2023).

A BN is defined by a directed acyclic graph (DAG) *G*, where each node in the graph represents a random variable and a set of parameters Θ that quantify the relationships between these variables. In a BN, an edge *X* → *Y* indicates that *X* is a direct parent of *Y*. This structure encodes conditional dependencies and independencies among variables. Given the DAG, the joint probability distribution of all variables factorises into the product of each variable’s conditional probability given its parents, capturing the strengths of the relationships between the variables (Kitson et al., 2023)

### Additive Bayesian network

Different methods and R packages can be used for BN construction. Initially, bnlearn() (Marco Scutari, 2024) was exploited to analyse network connections, but ultimately, an ABN approach with the abn() package (Furrer, R. et al., 2023) was chosen. ABN extends the generalised linear model (GLM) framework by factoring the joint probability distribution of all variables into a DAG, allowing both continuous and discrete variables to be simultaneously modelled. Unlike standard GLMs that predict only a single outcome, ABN captures complex dependencies among multiple variables (Delucchi et al., 2024), (Pittavino et al., 2017).

ABN learning involves structure and parameter learning (Furrer, R. et al., 2023). Given the multivariate nature of the present dataset, the Maximum Likelihood Estimation (MLE) approach was applied to estimate the dependencies and create the DAG. Structure learning employs the most probable explanation, a special case of maximum a posteriori estimation focusing on the most probable overall configuration of unobserved variables (Butz et al., 2018). Parameter estimation for Gaussian, Bernoulli, and Poisson nodes uses iterative reweighted least squares, whereas multinomial variables rely on a softmax-based conditional likelihood (Furrer, R. et al., 2023). In order to interpret the ABN, the multinomial variables are renamed to ensure the correct baseline category (e.g. ‘normal’ or ‘none’) for MLE scoring. Once the network is fitted, the model returns regression coefficients as posterior marginals. For Gaussian nodes, these are correlation coefficients on the standard linear scale, whereas for Poisson nodes, they represent log-rate ratios and for binomial or multinomial nodes, they are given on the log-odds scale (Furrer, R. et al., 2023). Gaussian, binomial, and multinomial nodes were used in this analysis.

The network’s fit was evaluated using AIC and BIC. AIC tends to select more complex models, whereas BIC, also known as the Schwarz information criterion or minimum description length, imposes a stronger penalty on complexity (Marco Scutari, 2024), (Jason Brownlee, 2020). For more heterogeneous data, like in this analysis, BIC often outperforms AIC, and given that BIC handles model complexity more economically, BIC is preferred for constructing ABN models (Brewer et al., 2016), (Ying et al., 2024). Therefore, BIC was used in this analysis to identify the optimal number of maximum parents and to fit the model. However, AIC and BIC both scale with the number of variables in the network, making comparisons of different networks and datasets, such as the delirium and HE cohorts, challenging (Kitson et al., 2023).

Arc strength was quantified via the average percentage of link strength, indicating the reduction in uncertainty for a given child variable when the states of its parents are known (Butz et al., 2018). To visualise the local structure around the variable of interest, the Markov blanket was used. The Markov blanket is the minimal set of influencing nodes independent of all other nodes. Therefore parents, children, and parents of children of the variable of interest are shown (Kitson et al., 2023).

Finally, because ABNs encode statistical rather than causal dependencies, arcs in these DAGs should not be interpreted as direct causal relationships (Pittavino et al., 2017). Ultimately, three separate data sets with defined variable distributions were used for the modelling. The delirium cohort and the DT cohort consist of the same variables and the same distribution, except for the ICD variable delirium and the ICD variable DT (see Table 4).

**Table 4:** Definition of variables/features and distribution used for modelling of delirium and DT cohort.

| Variable | Distribution | Definition |
| --- | --- | --- |
| **Sex** | binomial | Patient's administrative sex (Male/Female). |
| **Age** | gaussian | Patient's age in years (continuous variable). |
| **Ca_status** | multinomial | Categorisation of calcium levels (normal/hypo/hyper/both). |
| **K_status** | multinomial | Categorisation of potassium levels (normal/hypo/hyper/both). |
| **Na_status** | multinomial | Categorisation of sodium levels (normal/hypo/hyper/both). |
| **alb_status** | binomial | Albumin status (low/normal). |
| **ICDSC_max** | multinomial | Maximum ICDSC category for delirium assessment (0–2, 3, >3) |
| **PCT** | binomial | Procalcitonin analysed (yes/no) |
| **ICD_Dementia** | binomial | Diagnosis code indicator for dementia (yes/no). |
| **SAPS** | gaussian | SAPS (Simplified Acute Physiology Score), a continuous severity score. |
| **MDSI** | multinomial | A multi-category index (categories 1-9 describing the type of disease or comorbidities). |
| **Analgesics** | binomial | Use of analgesic medication (yes/no). |
| **Anaesthetics** | binomial | Use of anaesthetic medication (yes/no). |
| **Antipsychotics** | binomial | Use of antipsychotic medication (yes/no). |
| **Hyperoxia** | multinomial | Categorisation of oxygen level above normal (none/mild/moderate/severe). |
| **Hypoxia** | multinomial | Categorisation of oxygen level below normal (none/mild/moderate/severe). |
| **ICD_Thyroid** | binomial | Diagnosis code indicator for thyroid disease (yes/no). |
| **ICD_Malnutrition** | binomial | Diagnosis code indicator for malnutrition (yes/no). |
| **ICD_Diabetes** | binomial | Diagnosis code indicator for diabetes mellitus (yes/no). |
| **Pain** | binomial | Indicator of reported pain (yes/no). |
| **Ventilation** | binomial | Whether the patient was mechanically ventilated (yes/no). |
| **ICD_Delirium \*** | binomial | Diagnosis code indicator for delirium (yes/no). |
| **ICD_DTs \*** | binomial | Diagnosis code indicator for delirium tremens (yes/no). |

\*Difference between the two tables

The HE cohort has fewer variables, but an additional ammonia variable (see Table 5). The variables that are also used in the other cohorts are not listed again in Table 5 and can be referred to in Table 4.

**Table 5:** Definition of variables/features and distribution used for modelling of HE cohort.

| Variable | Distribution | Definition |
| --- | --- | --- |
| **Sex** | binomial | Patient's administrative sex (e.g., Male/Female). |
| **Age** | gaussian | Patient's age in years (continuous variable). |
| **Ca_status** | multinomial | Categorisation of calcium levels (normal/hypo/hyper/both). |
| **K_status** | multinomial | Categorisation of potassium levels (normal/hypo/hyper/both). |
| **Na_status** | multinomial | Categorisation of sodium levels (normal/hypo/hyper/both). |
| **Amm** | multinomial | Categorisation of ammonia levels (normal/hypo/hyper/both). |
| **ICDSC_max** | multinomial | Maximum ICDSC category for delirium assessment (e.g., 0–2, 3, >3). |
| **PCT** | binomial | Procalcitonin analysed (yes/no) |
| **SAPS** | gaussian | SAPS (Simplified Acute Physiology Score), a continuous severity score. |
| **Analgesics** | binomial | Use of analgesic medication (yes/no). |
| **Anaesthetics** | binomial | Use of anaesthetic medication (yes/no). |
| **Antipsychotics** | binomial | Use of antipsychotic medication (yes/no). |
| **Hyperoxia** | multinomial | Categorisation of oxygen level above normal (none/mild/moderate/severe). |
| **Hypoxia** | multinomial | Categorisation of oxygen level below normal (none/mild/moderate/severe). |
| **Ventilation** | binomial | Whether the patient was mechanically ventilated (yes/no). |
| **ICD HE** | Binomial | Diagnosis of hepatic encephalopathy (yes/no). |

### White- and Blacklisting

In BNs, constraints can incorporate domain knowledge into the structure learning process, helping to improve performance and counteract noise. These constraints act as requirements that the learnt structure must conform to (Kitson et al., 2023). In the present cohorts, only a conservative set of arcs was definitively blacklisted or whitelisted. For instance, demographic variables, such as age and sex, cannot be influenced by other measured factors and thus are blacklisted as child nodes. Other potential blacklists, such as medication-to-diagnosis paths, were tested but ultimately not enforced, as ongoing research has yet to confirm them as hard constraints.

Medication can influence appetite and thereby affect malnutrition (D’Alessandro et al., 2022). Malnutrition can reduce albumin levels (AMBOSS GmbH, 2024c). Certain medications can alter macronutrient intake in patients; however, none of the medications used in these cohorts are documented as having this effect (Prescott et al., 2018) But the contrary has also not been proven. Adverse drug reactions may lead to appetite loss and subsequent malnutrition or nutrient deficiencies (Kuzuya, 2023). Additionally, one systematic review found that reduced pulmonary function at baseline is associated with a higher risk of developing diabetes (Zhang et al., 2021). Some reviews suggest a possible causal link between certain anaesthetics and type-2 diabetes (Liu et al., 2023), and another study indicated that sepsis can raise blood glucose levels, thereby increasing the risk of type-2 diabetes (Gornik et al., 2010). No significant correlation has been demonstrated between thyroid hormones and various blood gas measurements—pH, PaO_2_, PaCO_2_, HCO_3_, or PaO_2_/FIO_2_—although the relationship between ventilation and thyroid function is not fully understood (Abdel Naby et al., 2015). The full blacklist of arcs can be found in the appendix, chapter 1.

Meanwhile, certain arcs are explicitly required. The SAPS II score is calculated using age and physiological variables like potassium, sodium among others (Afessa et al., 2007). Consequently, the relationships between potassium, sodium, and age with SAPS II were whitelisted to ensure these known relationships are preserved.

## Results

The results are derived from the characteristics of the patient cohort and the 3 ABN models and their comparison.

### Patient cohort characteristics

As explained in chapter 2.1.4, three cohorts were defined, which have deliriant symptoms and always include a control group. The variables are summarised in Table 6. The numerical variables show median and interquartile ranges. Categorical variables are listed with their values and percentages.

**Table 6:** Statistics of the three cohorts.

|  | **Delirium cohort \***<br>**n = 1378** | **Delirium Tremens co-**<br>**hort \*\***<br>**n = 892** | **Hepatic encephalopa-**<br>**thy cohort \*\*\***<br>**n = 251** |
| --- | --- | --- | --- |
| **Age, median, yr** | 66 (20-100) | 63 (21-97) | 62 (23-96) |
| **Sex, M, n (%)** | 904 (65.6%) | 550 (61.7%) | 140 (55%) |
| **Calcium** | 0: 529 (38.4%)<br>1: 767 (55.6%)<br>2: 69 (5.0%)<br>3: 14 (1.0%) | 0: 345 (38.7%)<br>1: 503 (56.4%)<br>2: 37 (4.1%)<br>3: 7 (0.8%) | 0: 58 (23.1%)<br>1: 161 (64.1%)<br>2: 43 (17.1%)<br>3: 4 (1.6%) |
| **Potassium** | 0: 746 (54.2%)<br>1: 260 (18.8%)<br>2: 352 (25.5%)<br>3: 20 (1.5%) | 0: 486 (54.5%)<br>1: 183 (20.5%)<br>2: 210 (23.5%)<br>3: 13 (1.5%) | 0: 144 (57.4%)<br>1: 60 (23.9%)<br>2: 43 (17.1%)<br>3: 4 (1.6%) |
| **Sodium** | 0: 704 (51.1%)<br>1: 131 (9.5%)<br>2: 538 (39.0%)<br>3: 5 (0.4%) | 0: 474 (53.1%)<br>1: 92 (10.3%)<br>2: 323 (36.2%)<br>3: 3 (0.3%) | 0: 121 (48.2%)<br>1: 43 (17.1%)<br>2: 85 (33.9%)<br>3: 2 (0.8%) |
| **Albumin** | 0: 21 (1.5%)<br>1: 1357 (98.5%) | 0: 17 (1.9%)<br>1: 875 (98.1%) | - |
| **Ammonia** | - | - | 0: 107 (42.6%)<br>1: 32 (12.7%)<br>2: 112 (44.6%) |
| **ICDSC** | 0-2: 687 (49.9%)<br>3: 153 (11.1%)<br>≥ 4: 538 (39.0%) | 0-2: 594 (66.6%)<br>3: 113 (12.7%)<br>≥ 4: 185 (20.7%) | 0-2: 139 (55.4%)<br>3: 25 (10.0%)<br>≥ 4: 87 (34.7%) |
| **PCT, 1, n (%)** | 1153 (83.7%) | 763 (85.5%) | 236 (94.4%) |
| **ICD Dementia, 1, n (%)** | 24 (1.7%) | 9 (1.0%) | - |
| **SAPS, median** | 49 (0-126) | 47 (2-126) | 53 (5-126) |
| **Mechanically venti-**<br>**lated, 1, n (%)** | 927 (67.2%) | 604 (67.9%) | 177 (70.5%) |
| **MDSI** | 1: 438 (31.8%)<br>2: 237 (17.2%)<br>3: 136 (9.9%)<br>4: 302 (21.9%)<br>6: 117 (8.5%)<br>8: 114 (8.3%) | 1: 252 (28.3%)<br>2: 163 (18.3%)<br>3: 90 (10.1%)<br>4: 221 (24.8%)<br>6: 69 (7.7%)<br>8: 76 (8.5%) | - |
| **Analgesics 1, n (%)** | 728 (52.8%) | 471 (52.8%) | 122 (48.6%) |
| **Anaesthetics, 1, n (%)** | 867 (62.9%) | 532 (59.6%) | 165 (65.7%) |
| **Antipsychotics, 1, n (%)** | 328 (23.8%) | 99 (11.2%) | 23 (9.2%) |
| **Hyperoxia** | None: 711 (51.6%)<br>Mild: 363 (26.3%)<br>Moderate: 186 (13.5%)<br>Severe: 118 (8.6%) | None: 464 (52.0%)<br>Mild: 230 (25.8%)<br>Moderate: 118 (13.2%)<br>Severe: 80 (9.0%) | None: 123 (49.0%)<br>Mild: 59 (23.5%)<br>Moderate: 39 (15.5%)<br>Severe: 30 (12.0%) |
| **Hypoxia** | None: 523 (38.0%)<br>Mild: 602 (43.7%)<br>Moderate: 146 (10.6%)<br>Severe: 107 (7.8%) | None: 373 (41.8%)<br>Mild: 377 (42.3%)<br>Moderate: 92 (10.3%)<br>Severe: 60 (6.7%) | None: 109 (43.4%)<br>Mild: 105 (41.8%)<br>Moderate: 28 (11.2%)<br>Severe: 9 (3.6%) |
| **ICD Thyroid, 1, n (%)** | 102 (7.4%) | 60 (6.7%) | - |
| **ICD Malnutrition, 1, n (%)** | 136 (9.9%) | 75 (8.4%) | - |
| **ICD Diabetes, 1, n (%)** | 297 (21.6%) | 160 (17.9%) | - |
| **Pain, 1, n (%)** | 494 (35.8%) | 325 (43.2%) | - |
| **ICD_Delirium** | 514 (37.3%) | - | - |
| **ICD_DTs** | - | 28 (3.1%) | - |
| **ICD HE** | - | - | 61 (24%) |

Definition of coding: 0 = normal, 1 = hypo, 2 = hyper, 3 = both,
MDSI: 1 = Cardiovascular, 2 = Respiratory and ENT, 3 = Gastrointestinal, 4 = Neurological, 5 = Metabolic-endocrinological, 6 = Trauma, 7 = Urogenital, 8 = Other, 9 = --
\* No delirium tremens (ICD F10.4) and no hepatic encephalopathy (ICD K72.7)
\*\* No delirium (ICD F05) and no hepatic encephalopathy (ICD K72.7)
\*\*\* No delirium tremens (ICD F10.4) and no delirium (ICD F05)

Overall, 880 patients showed no delirium according to the ICDSC score, whereas 552 had delirium based on the F05 ICD code, and 620 were categorised as delirious by the ICDSC. In the DT cohort, 28 patients had DT defined by ICD code, but 185 patients had an ICDSC score of 4 or higher.

Across all cohorts, hypocalcaemia emerged as the most common calcium imbalance. Potassium levels were typically normal, but patients in the delirium and DT cohorts tended to have hyperkalaemia more often. In contrast, those in the HE cohort were more likely to exhibit hypokalaemia. In terms of albumin levels, low albumin was prevalent in the cohorts at over 98%, which aligns with existing research indicating frequent hypoalbuminemia in critically ill patients (Vincent et al., 2003). Most patients in the delirium and DT cohorts were admitted to the ICU for cardiovascular or neurological conditions. Notably, a higher proportion of patients in the delirium cohort (23.8%) received antipsychotic medications compared to patients in the other cohorts. Finally, hypoxia was more common overall than hyperoxia.

To further analyse the delirium cohort, which is the focus of this study, Table 7 compares the variables of cases without delirium based on an ICDSC score of less than 3, threshold cases based on an ICDSC score of 3, and delirium cases based on an ICDSC score of 4 and higher. Currently, an ICDSC score of at least 4 is required to diagnose delirium. However, it is being discussed whether patients with an ICDSC score of 3 should also be considered delirious (‘Screening for Delirium with the Intensive Care Delirium Screening Checklist (ICDSC)’, 2018).

**Table 7:**
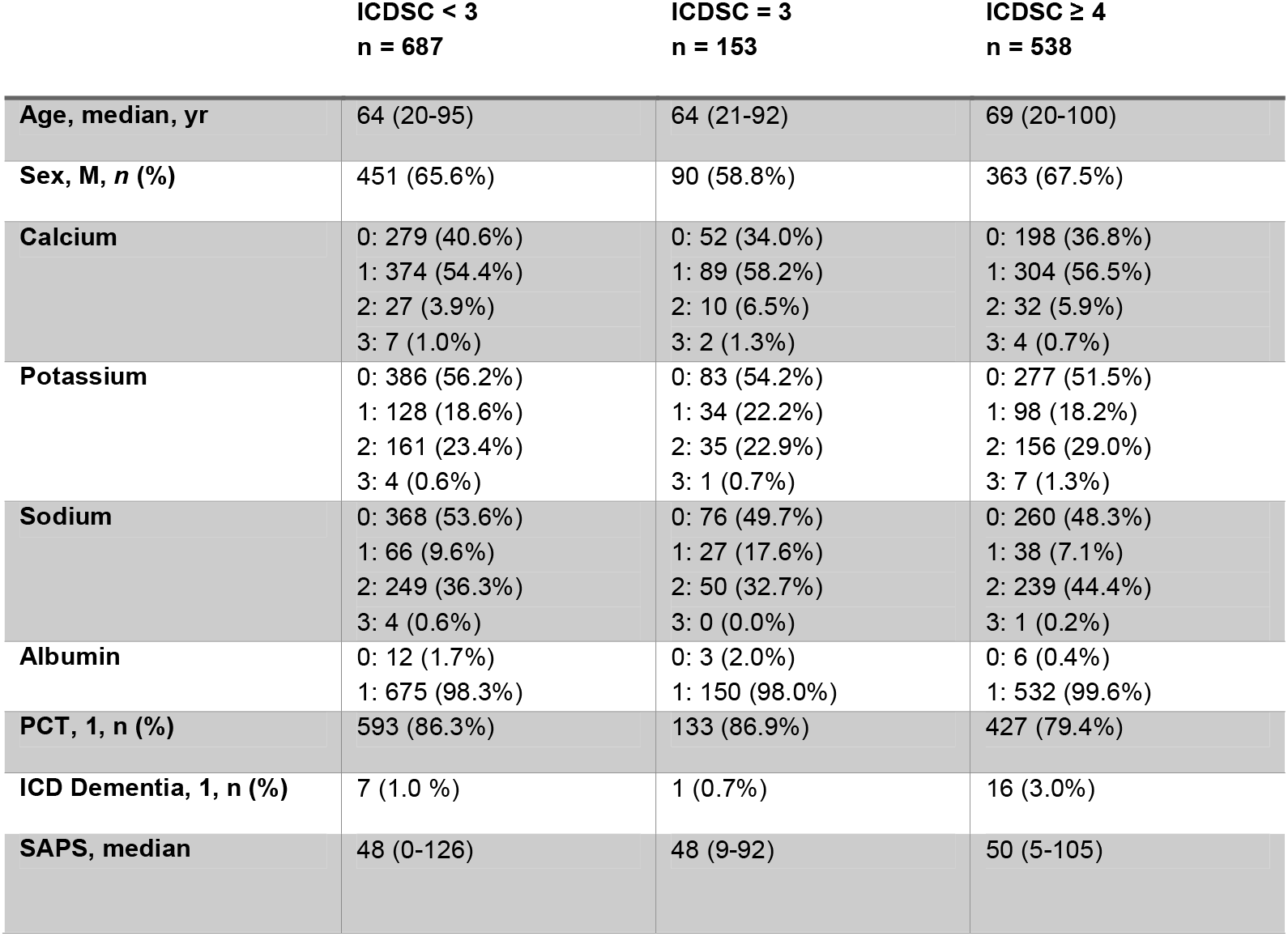
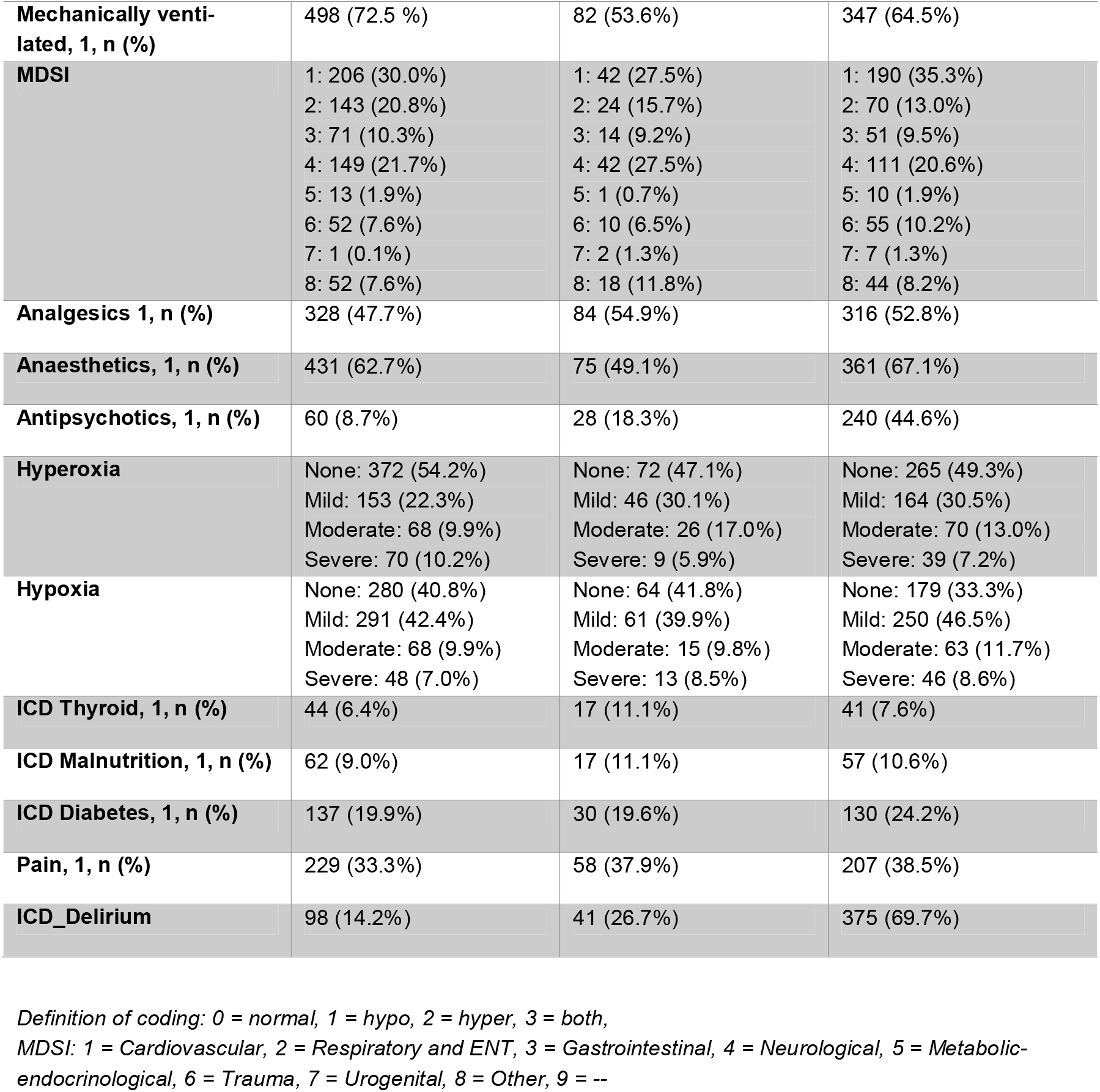
Statistics of the delirium cohort.

|  | **ICDSC < 3**<br>**n = 687** | **ICDSC = 3**<br>**n = 153** | **ICDSC ≥ 4**<br>**n = 538** |
| --- | --- | --- | --- |
| **Age, median, yr** | 64 (20-95) | 64 (21-92) | 69 (20-100) |
| **Sex, M, n (%)** | 451 (65.6%) | 90 (58.8%) | 363 (67.5%) |
| **Calcium** | 0: 279 (40.6%)<br>1: 374 (54.4%)<br>2: 27 (3.9%)<br>3: 7 (1.0%) | 0: 52 (34.0%)<br>1: 89 (58.2%)<br>2: 10 (6.5%)<br>3: 2 (1.3%) | 0: 198 (36.8%)<br>1: 304 (56.5%)<br>2: 32 (5.9%)<br>3: 4 (0.7%) |
| **Potassium** | 0: 386 (56.2%)<br>1: 128 (18.6%)<br>2: 161 (23.4%)<br>3: 4 (0.6%) | 0: 83 (54.2%)<br>1: 34 (22.2%)<br>2: 35 (22.9%)<br>3: 1 (0.7%) | 0: 277 (51.5%)<br>1: 98 (18.2%)<br>2: 156 (29.0%)<br>3: 7 (1.3%) |
| **Sodium** | 0: 368 (53.6%)<br>1: 66 (9.6%)<br>2: 249 (36.3%)<br>3: 4 (0.6%) | 0: 76 (49.7%)<br>1: 27 (17.6%)<br>2: 50 (32.7%)<br>3: 0 (0.0%) | 0: 260 (48.3%)<br>1: 38 (7.1%)<br>2: 239 (44.4%)<br>3: 1 (0.2%) |
| **Albumin** | 0: 12 (1.7%)<br>1: 675 (98.3%) | 0: 3 (2.0%)<br>1: 150 (98.0%) | 0: 6 (0.4%)<br>1: 532 (99.6%) |
| **PCT, 1, n (%)** | 593 (86.3%) | 133 (86.9%) | 427 (79.4%) |
| **ICD Dementia, 1, n (%)** | 7 (1.0 %) | 1 (0.7%) | 16 (3.0%) |
| **SAPS, median** | 48 (0-126) | 48 (9-92) | 50 (5-105) |
| **Mechanically venti-**<br>**lated, 1, n (%)** | 498 (72.5 %) | 82 (53.6%) | 347 (64.5%) |
| **MDSI** | 1: 206 (30.0%)<br>2: 143 (20.8%)<br>3: 71 (10.3%)<br>4: 149 (21.7%)<br>5: 13 (1.9%)<br>6: 52 (7.6%)<br>7: 1 (0.1%)<br>8: 52 (7.6%) | 1: 42 (27.5%)<br>2: 24 (15.7%)<br>3: 14 (9.2%)<br>4: 42 (27.5%)<br>5: 1 (0.7%)<br>6: 10 (6.5%)<br>7: 2 (1.3%)<br>8: 18 (11.8%) | 1: 190 (35.3%)<br>2: 70 (13.0%)<br>3: 51 (9.5%)<br>4: 111 (20.6%)<br>5: 10 (1.9%)<br>6: 55 (10.2%)<br>7: 7 (1.3%)<br>8: 44 (8.2%) |
| **Analgesics 1, n (%)** | 328 (47.7%) | 84 (54.9%) | 316 (52.8%) |
| **Anaesthetics, 1, n (%)** | 431 (62.7%) | 75 (49.1%) | 361 (67.1%) |
| **Antipsychotics, 1, n (%)** | 60 (8.7%) | 28 (18.3%) | 240 (44.6%) |
| **Hyperoxia** | None: 372 (54.2%)<br>Mild: 153 (22.3%)<br>Moderate: 68 (9.9%)<br>Severe: 70 (10.2%) | None: 72 (47.1%)<br>Mild: 46 (30.1%)<br>Moderate: 26 (17.0%)<br>Severe: 9 (5.9%) | None: 265 (49.3%)<br>Mild: 164 (30.5%)<br>Moderate: 70 (13.0%)<br>Severe: 39 (7.2%) |
| **Hypoxia** | None: 280 (40.8%)<br>Mild: 291 (42.4%)<br>Moderate: 68 (9.9%)<br>Severe: 48 (7.0%) | None: 64 (41.8%)<br>Mild: 61 (39.9%)<br>Moderate: 15 (9.8%)<br>Severe: 13 (8.5%) | None: 179 (33.3%)<br>Mild: 250 (46.5%)<br>Moderate: 63 (11.7%)<br>Severe: 46 (8.6%) |
| **ICD Thyroid, 1, n (%)** | 44 (6.4%) | 17 (11.1%) | 41 (7.6%) |
| **ICD Malnutrition, 1, n (%)** | 62 (9.0%) | 17 (11.1%) | 57 (10.6%) |
| **ICD Diabetes, 1, n (%)** | 137 (19.9%) | 30 (19.6%) | 130 (24.2%) |
| **Pain, 1, n (%)** | 229 (33.3%) | 58 (37.9%) | 207 (38.5%) |
| **ICD_Delirium** | 98 (14.2%) | 41 (26.7%) | 375 (69.7%) |

Definition of coding: 0 = normal, 1 = hypo, 2 = hyper, 3 = both,
MDSI: 1 = Cardiovascular, 2 = Respiratory and ENT, 3 = Gastrointestinal, 4 = Neurological, 5 = Metabolic-endocrinological, 6 = Trauma, 7 = Urogenital, 8 = Other, 9 = --

In patients with an ICDSC score of 4 or higher, the median age (69 years) was greater than among those with ICDSC < 3 (64 years) or ICDSC = 3 (64 years). This group also had a slightly higher proportion of male patients, with 67.5% compared to 65% in ICDSC > 3 and 58% in ICDSC = 3. In line with the literature, this suggests that older male patients are especially prone to delirium. Many of these ICDSC ≥ 4 patients are treated in the ICU for cardiovascular diseases or trauma, whereas respiratory or ENT-related causes appear less frequently than those in the ICDSC < 4 groups. Interestingly, mechanical ventilation was most common in patients with ICDSC < 3 (72.5%), followed by ICDSC ≥ 4 (64.5%), and least frequent in ICDSC = 3 (53.6%), which does not indicate that delirium patients are more frequently ventilated, as the literature states ((Qureshi & Arthur, 2023), (AMBOSS GmbH, n.d.)). Examining lab values revealed that normal potassium and sodium ranges were somewhat less frequent in the ICDSC ≥ 4 group, with a higher portion of hyperkalaemia and hypernatremia. A study showed that 33.0% of patients with delirium had hyponatraemia and 15.1% had hypernatraemia (Tran et al., 2021). In contrast, in this cohort, 7% of the delirium patients had hyponatraemia, and 44% had hypernatremia.

More patients with ICDSC = 3 (52.9%) and ICDSC ≥ 4 (50.7%) experienced hyperoxia, especially the mild and moderate variation. More patients with ICDSC ≥ 4 experienced hypoxia, which agrees with the literature (Qureshi & Arthur, 2023), (AMBOSS GmbH, n.d.),. The SAPS score was quite close through all three groups, but the highest in the ICDSC ≥ 4 group with a median score of 50. Most patients across all groups had low albumin levels, reflecting typical ICU findings (Tran et al., 2021), (Vincent et al., 2003).

In terms of therapy, a markedly higher proportion of ICDSC ≥ 4 patients received antipsychotics (44.6%) compared to those in the ICDSC = 3 (18.3%) and ICDSC < 3 (8.7%) groups. Olanzapine and olanzapine were most often administered (see Figure 3), with only a few patients receiving haloperidol and risperidone, the ranges of these medications being highest at an ICDSC of 4. Patients with an ICDSC of 6 receiving the most medications overall.

**Figure 3:**
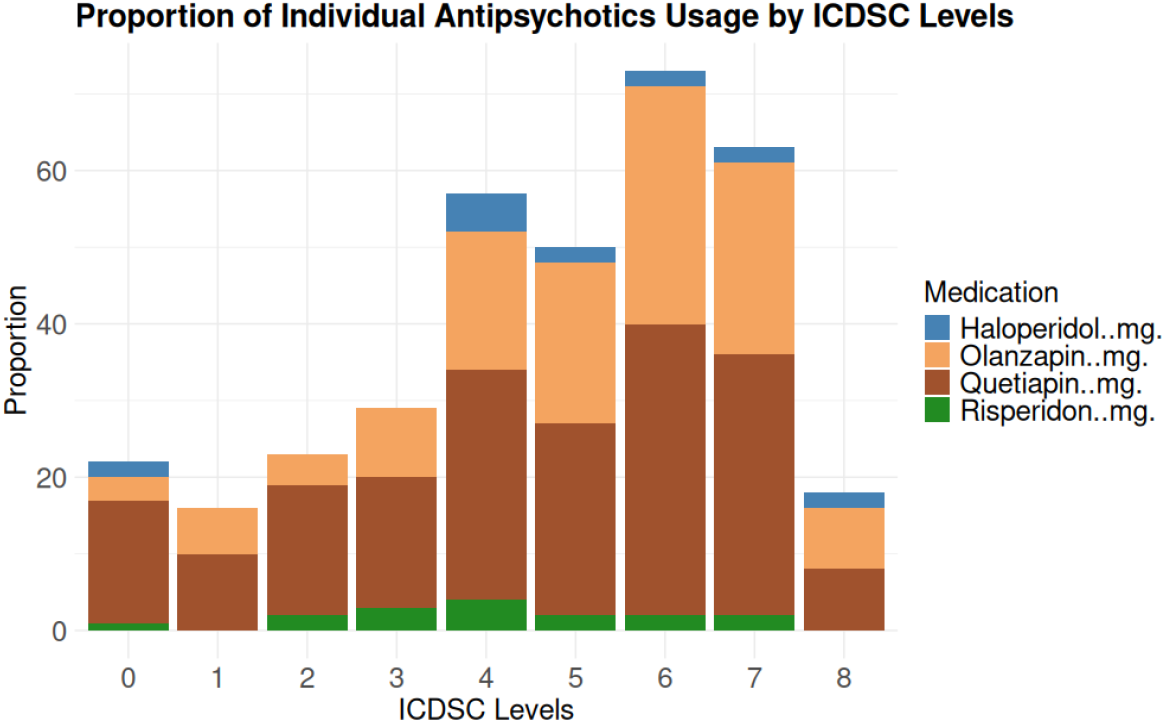
Antipsychotics administration by ICDSC levels

Although analgesic use was somewhat elevated in the ICDSC = 3 group (54.9%), it remained substantial across all three categories, with 47.7% in ICDSC < 3 and 52.8% in ICDSC ≥ 4. Not only are the analgesics an influential factor on delirium but also the pain managed with it (Duprey et al., 2021). This is consistent with the statistics of this cohort. The highest percentage of patients reporting pain were in the ICDSC ≥ 4 group (38.5%), closely followed by the ICDSC = 3 group (37.9%).

All three ICDSC groups also had patients with an ICD-based delirium diagnosis, potentially reflecting either inaccuracies in documentation or a delirium that occurred outside of the ICU setting for the group ICDSC < 3. Notably, the proportion of patients with an ICD delirium diagnosis increased as the ICDSC score increased with 14.2% in ICDSC < 3, 26.8% in ICDSC = 3, and 69.7% in ICDSC ≥ 4.

### Additive Bayesian network

Since two of three cohorts have more than the recommended maximum of 20 variables, the calculation was very time- and memory-intensive. For the delirium and delirium tremens cohorts, it was not possible to achieve a maximum number of parents of more than 7; the time required for other counts of maximum parents can be seen in the Appendix, chapter 6. However, the BIC has not yet reached its plateau. Calculating the ABN of the delirium cohort and the delirium tremens cohort with a maximum of 7 parents took just over 3 days. The HE cohort comprised 250 patients and 16 parameters, resulting in a lower computational effort relative to that of the larger cohorts. Although the AIC and BIC metrics began to plateau at approximately 10 parents (see Appendix chapter 6), a maximum of 15 parents were tested. Using three cores, the score cache required approximately 15 min to build, followed by an additional 10 minutes to compute the DAG.

### ABN Delirium cohort

Since ICDSC is modelled as a multinomial variable, its links (see Figure 4) should be interpreted on a log-odds basis. A notable connection between ICDSC and ICD delirium was found, which aligns with expectations that both variables capture a form of delirium. The association between ICDSC ≥4 and ICD Delirium was insignificant but as expected, lower ICDSC scores have a negative association with delirium. However, as noted, ICDSC is considered more reliable in this context. The model identified Age and Ventilation as parents of the ICDSC values; interestingly, Ventilation increased the odds of having an ICDSC of 3, whereas Age and Ventilation were both associated with a lower probability of ICDSC ≥4 than ICDSC <3. Ventilation reduces the odds of having an ICDSC score ≥4 by approximately 33.4% and a one-year increase in age decreases the log-odds of having an ICDSC score ≥4 by 0.82. This finding contradicts some of the literature, as both age and ventilation are typically seen as risk factors for delirium.

**Figure 4:**
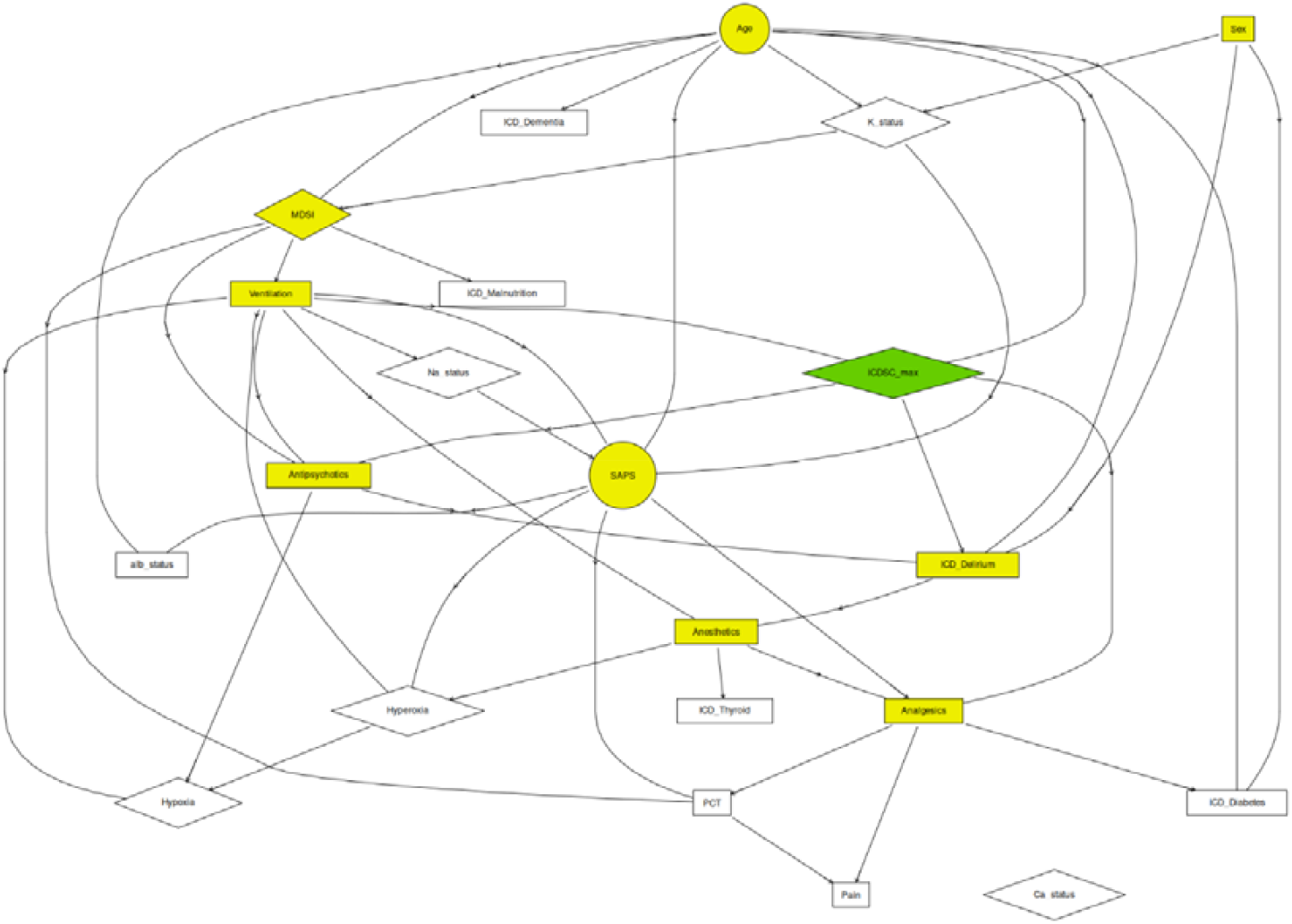
ABN of the delirium cohort with the variables of the Markov blanket in yellow for the variable ICDSC_max in green

Further child nodes of ICDSC include Antipsychotics, ICD Delirium, and Analgesics. Greater ICDSC values increased the likelihood of antipsychotic usage, whereas lower ICDSC values were associated with decreased probabilities of ICD delirium and a stronger negative association with analgesics. That means that a higher ICDSC value is less likely to result in analgesic administration.

ICD Delirium itself had additional parents— Antipsychotics, Sex, and Age. Male patients, older patients, and those given antipsychotics all demonstrated increased log-odds of having an ICD Delirium code, which aligns closely with existing literature. Specifically, patients receiving antipsychotics are almost four times more likely to experience ICD delirium than patients not receiving antipsychotics, and men are about 87% more likely to experience ICD delirium than women. ICD delirium was also associated with anaesthesia, patients diagnosed with delirium were more likely to receive anaesthesia.

Interestingly, as Ventilation is an important factor MDSI 2 (Respiratory and ENT) and MDSI 6 (Trauma) seemed to increase the probability of ventilation, whereas only MDSI 7 (Urogenital) reduces the odds—both in comparison to the baseline MDSI 1 (Cardiovascular). Regarding Analgesics and ICD Diabetes, the direction of influence is somewhat unclear, but the association remains in the model because no definitive evidence warranted blacklisting this path. Certain MDSI categories, particularly 4 (Neurological) and 5 (Metabolic-Endocrinological), demonstrate a negative association with Antipsychotics, and ventilation also lowered the probability of receiving antipsychotics. Higher SAPS scores reduced the probability of analgesic usage, whereas anaesthetics increased it. Antipsychotics increased the likelihood of mild hyperoxia but lowered the probability of moderate or severe hypoxia, and ventilation reduced the chance of hypoxia except in severe cases. Severe hyperoxia demonstrated a strong positive interaction with severe hypoxia, whereas mild, moderate, and severe hyperoxia reduced the probability of moderate hypoxia.

Finally, PCT—an infection marker—exhibited a positive association with pain, reflecting that elevated infection risk often correlates with painful inflammatory processes. Moreover, the use of analgesics also increases the odds of pain.

Especially strong were the arcs from the parents to SAPS, followed by the arcs to MDSI and ICD Delirium. The arcs between Potassium and MDSI exhibit the largest standard error, indicating a high degree of variability.

### ABN Delirium Tremens cohort

The ICD DT is modelled as a binomial variable (Bernoulli), meaning that the correlations should be interpreted on a log-odds basis (see Figure 5). The ABN revealed a clear relationship between ICDSC and ICD DT, which is consistent with both variables indicating delirium. However, as previously discussed, ICDSC is considered the more reliable measure, whereas ICD DT specifically captures only one subtype of delirium. In this cohort, ICD DT was the focus, as ICDSC does not distinguish delirium types.

**Figure 5:**
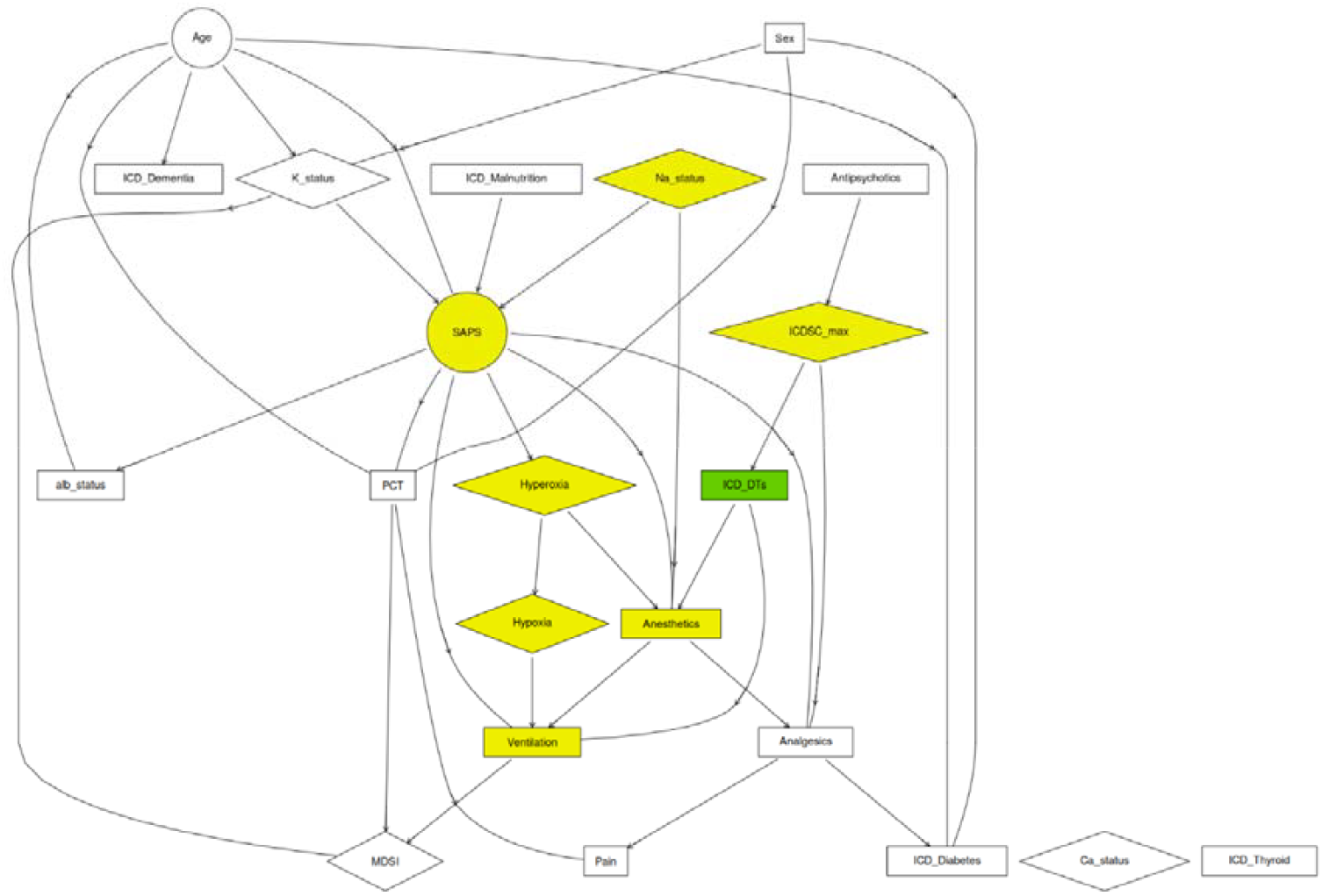
ABN of the delirium tremens cohort with the variables of the Markov blanket in yellow for the variable ICDSC_max in green

According to the model, ICD DT is influenced solely by the ICDSC score, which in turn is closely associated with antipsychotic administration. A higher ICDSC score increases the likelihood of DT. ICDSC itself was influenced by Antipsychotics, especially for scores ≥4. Administration of antipsychotics increases the log-odds of having ICDSC ≥4 by 1.86 compared to ICDSC <3. Furthermore, a higher ICDSC ≥4 was associated with a greater likelihood of analgesic use, whereas a lower ICDSC ≤3 reduces that likelihood. Once DT occurs, it strongly increases the probability of receiving anaesthetics while decreasing the probability of ventilation. Meanwhile, ICDSC scores of 4 or above meant that analgesics were more likely to be administered; ICDSC 3 or below revealed a much lower probability of analgesic use. Anaesthetics were more likely to be given if both hyper- and hypocalcaemia, severe hyperoxia, or DT occurred.

This cohort lacked data on alcohol consumption, but it broadly served as a comparison group to the primary delirium cohort. The link between ICDSC and delirium in this group mirrors patterns seen elsewhere, although Ventilation and certain MDSI parameters appear as child nodes here rather than parent nodes. Overall, the model’s structure can be considered similar to that of the primary cohort.

### ABN Hepatic Encephalopathy cohort

As the target variable ‘ICD HE’ is modelled as a binomial variable (Bernoulli), its links should be interpreted on a log-odds basis (see Figure 6). An ICDSC value of 0–2 demonstrated a stronger negative association with HE than ICDSC 3 or ≥4, indicating that lower ICDSC scores reduce the likelihood of HE. High ammonia levels (>47 μmol/L) raised the log odds of HE by 0.74, whereas low ammonia (<23 μmol/L) lowered it. The ICDSC score was influenced by antipsychotic use, and these drugs increased the chance of scoring ≥4 while substantially reducing the chance of scoring exactly 3.

**Figure 6:**
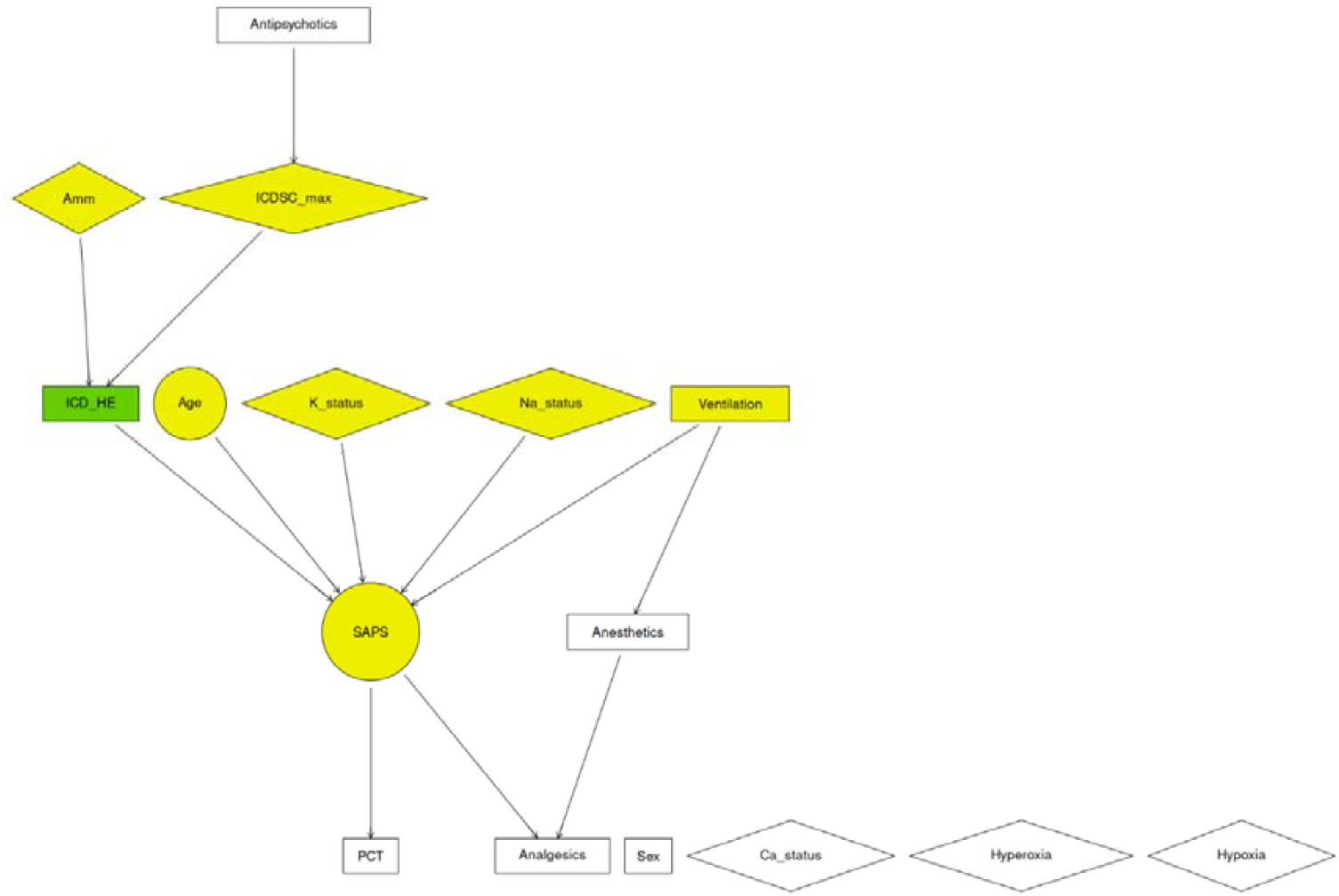
ABN of the Hepatic Encephalopathy cohort with the variables of the Markov blanket in yellow for the variable ICD_HE in green

Age, sodium status, potassium status, ventilation status, and HE are all strongly linked to SAPS, which means the uncertainty for the SAPS is low, given these parents. Hyponatraemia and hypokalaemia are associated with lower SAPS values, whereas older age and the presence of HE both increase SAPS, potentially indicating a higher risk of ICU mortality. The model also confirms a logical connection between anaesthetics and ventilation, reflecting the routine clinical practice of ventilated patients typically receiving sedation or anaesthesia.

The model shows no links to and from the variables Gender, Calcium status, Hyperoxia, and Hypoxia.

### Comparison of the three cohorts

Across the three cohorts, ventilation played varying roles in the onset and progression of delirium-related conditions. In the Delirium cohort, ventilation is a parent of ICDSC and ICD Delirium, whereas in DT, the direction is reversed — DT lowered the likelihood of ventilation rather than being influenced by it. In the HE cohort, Ventilation interacted primarily with the SAPS score. Likewise, MDSI categories (neurological and metabolic) are parents of the Ventilation or Antipsychotic variables in the delirium and HE cohort but appeared as child nodes in DT. The variable Ventilation was introduced later into the analysis, and it shows that hyperoxia and hy-poxia were largely overshadowed by ventilation, it supersedes oxygenation parameters as a more powerful predictor. Anaesthetics were more likely to be administered if severe hyperoxia or DT occurred, which agrees with the delirium cohort, where delirium and ventilation increased the likelihood of receiving anaesthetics. Moreover, the likelihood of anaesthetics increased in the HE cohort if the patient was on mechanical ventilation.

A notable pattern emerged between the ICDSC score and Antipsychotics. In the delirium cohort, higher ICDSC values tended to increase the possibility of antipsychotic administration, which was then associated with the ICD diagnosis of delirium. In the HE cohort, the use of antipsychotics increased the ICDSC value. In DT, however, DT itself stems from an elevated ICDSC, and that high score was also associated with greater antipsychotic usage.

Several observations remained consistent across models. Overall, Sodium, Potassium, Age, and Ventilation were key variables for SAPS in all cohorts. The arcs between PCT, infection, and analgesics are the same in the two delirium cohorts that have the pain variable. SAPS is associated with less likelihood of analgesic administration and higher likelihood of having procalcitonin tested and therefore infection in all 3 cohorts.

## Discussion

### Interpretation of results

The delirium cohort model revealed that mechanical ventilation increased the probability of having an ICDSC of 3, whereas both age and ventilation lowered the probability of ICDSC ≥4 in comparison to ICDSC <3. This finding contradicts the literature, as both age and ventilation are typically seen as risk factors for delirium (Mann, 2018), (Qureshi & Arthur, 2023), (Ali & Cascella, 2024), (AMBOSS GmbH, n.d.). Interestingly, the statistics of the data do not correlate with the results of the model, as can be seen in Table 7 and Figure 7: patients with ICDSC ≥4 had a higher median age (69 years) than those scoring <3 or exactly 3 (both 64 years), and mechanical ventilation was most frequent among patients with ICDSC <3 (72.5%), followed by ICDSC ≥4 (64.5%), and finally ICDSC =3 (53.6%). The negative association between ventilation and ICDSC ≥4 could stem from the fact that even though ICDSC can be measured in intubated patients, nurses often reported that delirium was difficult to evaluate in intubated patients (Mart et al., 2021). While age is typically positively associated with delirium, the negative coefficient for age in the model for ICDSC ≥4 needs to be further evaluated. Several factors could explain this result, including sampling bias, but it is not as likely as the data characteristics suggesting something else. Unmodeled interactions between age and other variables, overfitting, or noise are more likely. Overfitting, particularly in complex models with numerous predictors and limited sample sizes in certain categories, is a recognised issue (Furrer, R. et al., 2023). Despite this, the arcs within the model remained consistent, with a maximum of four and eight parent nodes. The possibility of unmodeled interactions requires further exploration. For instance, research suggests that age may not significantly impact delirium, and younger patients with acute heart failure may exhibit increased susceptibility to delirium due to brain hypoperfusion with higher ejection fraction (Han et al., 2022). Similarly, another study reported no difference in delirium outcomes across age groups but noted that younger patients were more frequently administered fentanyl, benzodiazepines, lorazepam, and midazolam (Guo et al., 2024).

**Figure 7:**
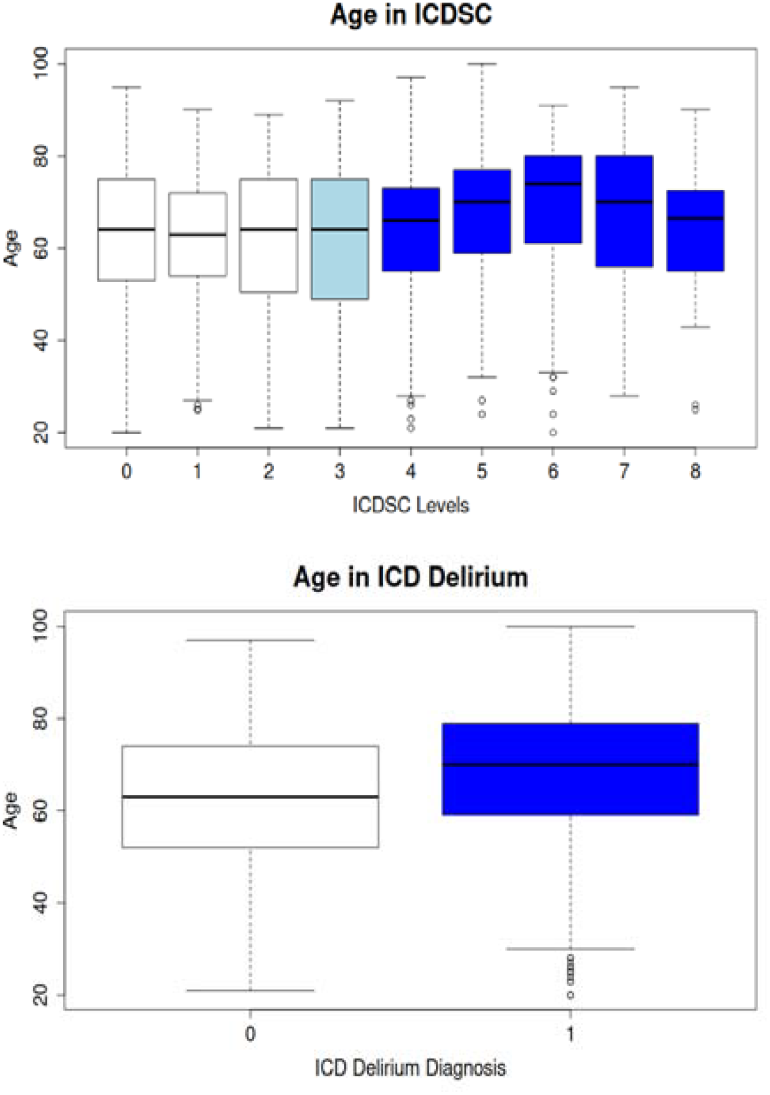
Age distributions stratified by ICDSC level and delirium diagnosis by ICD code

The model identified interesting relationships between age, MDSI, ventilation, antipsychotics, and ICD delirium. Specific MDSI categories (1, 3, and 6—cardiovascular, respiratory, ENT, and trauma) were associated with an increased likelihood of ventilation, while MDSI 7 (urogenital) reduced the likelihood of ventilation. Ventilation was found to decrease the likelihood of antipsychotic use. Higher ICDSC scores were positively associated with antipsychotic administration, which in turn increased the likelihood of delirium. Antipsychotics demonstrated a strong positive association with delirium diagnoses in all three cohorts. Nonetheless, it remains unclear whether antipsychotics contribute to delirium or are administered pre-emptively in response to observed delirium symptoms. Analysis of the raw data indicated that while antipsychotics were given both prior to and after the delirium measurement, mostly in a 10-hour frame, a slightly bigger part were given shortly before the ICDSC measurement. Clinicians often use antipsychotics to manage agitation and maintain patient and staff safety, especially under high workload pressures, even though this practice frequently falls outside evidence-based guidelines. While antipsychotics can alleviate agitation and psychotic symptoms, especially in patients with hyperactive delirium due to their sedative effects, they do not address the underlying pathology of delirium and may prolong its duration (Tomlinson et al., 2024). However, in delirium tremens, the use of benzodiazepines, clomethiazole, haloperidol, and clonidine for treatment is recommended (Kukolja & Kuhn, 2021).

This ABN analysis found no significant influence of dementia, malnutrition, or multiple comorbidities (measured via SAPS) on delirium, which contradicts previous literature (Mann, 2018), (Qureshi & Arthur, 2023), (Ali & Cascella, 2024), (AMBOSS GmbH, n.d.). However, the statistical data supported the literature, as patients with ICDSC scores >3 were more likely to have dementia than those with ICDSC scores <4. A study states that severe pain was inversely associated with transitioning to delirium. And opioid administration in awake, non-delirious patients increased the risk of delirium the following day (Duprey et al., 2021). However, this model did not find analgesics or pain to significantly influence delirium. The statistics of the data in Table 7 suggest a correlation between hyperkalaemia and hypernatraemia with delirium, but in the ABN no association was found. Instead, ventilation, antipsychotics, age, and sex emerged as the most significant variables.

The ABN demonstrates its reliability as a model by accurately identifying established relationships and influences. It highlights the strong connection between SAPS and its known and whitelisted predictors sodium, potassium, and age (Afessa et al., 2007). In addition, ventilation, malnutrition, and hepatic encephalopathy are associated with a higher SAPS score. The link between ventilation and SAPS is reasonable, as PaO_2_ values and FiO_2_ values are used to calculate the SAPS score; the malnutrition and HE parameters are new, and it should be questioned whether this arc is actually in the direction of SAPS or whether it should be the other way round. In the DT cohort, an increased likelihood of ventilation was also associated with the occurrence of delirium tremens. These findings align with existing literature, which reports that delirium in intensive care patients is linked to a higher probability of mechanical ventilation and a threefold increase in mortality risk (Hermes et al., 2022). Furthermore, the ABN correctly illustrates the correlation between pain, infection, and analgesic use and confirms the recognised association be-tween elevated ammonia levels and the development of HE (Aldridge et al., 2015).

The hypothesis that previously unrecognised connections influence delirium is partially supported by the findings. While several known associations, such as age and ventilation, were confirmed, the identification of reduced delirium likelihood (ICDSC ≥4) associated with age and ventilation was novel. The connection between antipsychotics and delirium is already discussed and not cleared up through this analysis. Conversely, all connections involving ICD delirium were consistent with expectations. This may be partly explained by the primary use of ICD codes for billing, which could result in delirium being coded primarily in cases requiring additional interventions such as ventilation or the administration of medication. The relationship between antipsychotics and delirium, while already a topic of discussion, remains unresolved in this analysis as there are arcs in both directions.

### Clinical implications

The relationship between antipsychotics and delirium is complex. This model highlights the bidirectional nature of this association: ICDSC scores of 4 and above increase the likelihood of antipsychotic use, but the use of antipsychotics, in turn, raises the probability of developing delirium and delirium tremens and heightens the ICDSC score in the HE cohort. Antipsychotics are frequently used as first-line treatment to manage the hyperactive form of delirium, though evidence supporting their efficacy for delirium treatment in both ICU and non-ICU settings remains limited (Kukolja & Kuhn, 2021).

The model developed in this study further identifies advanced age with a lower likelihood of delirium identified by ICDSC. This finding aligns with existing literature suggesting the need for greater inclusion of younger patients in delirium research. Studies on postoperative delirium have demonstrated that younger patients face a comparable risk of delirium to that of older individuals (Han et al., 2022), (Guo et al., 2024).

Additionally, the use of an ABN presents a valuable methodological approach for examining delirium and other conditions characterised by significant uncertainty. The robustness of connections between variables across all three cohorts in this study highlights the utility of this method. Furthermore, the HE cohort model corroborates existing evidence that elevated ammonia levels are strongly associated with a higher likelihood of HE (Aldridge et al., 2015).

## Limitations

A limitation of this analysis is the small dataset. Additional variables known to influence delirium, such as glucose levels, urinary retention, operation type and length, alcohol and nicotine consumption, and sensory impairments, could not be analysed due to missing data and the need to limit the total number of variables.

There are limitations for the main target variable. The ICDSC measurements, recorded at least once per shift by nursing staff, are considered highly reliable. In contrast, ICD codes are extracted from medical reports primarily designed for billing and diagnosis tracking. These reports, often written after extended hospital stays, can overlook conditions like delirium if overshadowed by more severe diagnoses or events. This is shown by the difference between the number of patients with ICDSC over 3 (620) and the number of patients with ICD delirium (552). This discrepancy highlights the importance of relying on ICDSC for a more accurate and consistent assessment of delirium. A proposed approach to improve delirium classification was to define ‘real delirium’ as ICD codes for delirium combined with ICDSC scores ≥4 while categorising ICDSC <4 and no diagnosis as ‘no delirium’. However, this would have significantly reduced the dataset and was deemed impractical for this analysis.

Increased blacklisting could improve the model’s clinical relevance. For instance, the arc from Ventilation or Potassium to MDSI may be clinically irrational and may be better reversed or excluded. Similarly, the arc from analgesics to diabetes likely represents the opposite causal relationship.

When interpreting the outcome labels, the risk of overfitting must be acknowledged, as this model does not adjust for it. The MLE DAG strategy can lead to spurious relationships, especially with small datasets, necessitating the inclusion of measures such as majority consensus networks that average over competing models to identify robust structural features. An alternative is also to prune the DAG with the highest score by crossing it with the DAG with majority consensus, retaining only the agreed features. Alternatively, methods like Bayesian model averaging or bootstrapping (using tools like JAGS or WinBUGS) can help identify robust features while addressing overfitting (Furrer, R. et al., 2023).

Furthermore, the computational burden must be considered. The mostProbable() function becomes both time- and memory-intensive as the number of nodes increases. Computational effort with more parameters is something to be incorporated in planning. While additive models can be analysed efficiently for networks of up to 20 nodes on most machines, beyond this size, the necessary computing resources and runtime often become impractical (Furrer, R. et al., 2023). Although additive models can often be handled efficiently for networks of up to approximately 20 nodes, resource requirements become impractical beyond that size. Even on a high-capacity server (250 GB RAM, 32 GPUs), the process remained inefficient with 22 nodes versus 14 nodes (see Appendix chapter 6). Therefore, the maximum number of parents per variable in the delirium and DT cohorts had to be set to 7. Future analyses should explore additional parents, as the BIC curve has not yet plateaued.

Finally, this analysis is restricted to patients in the ICU, a setting already recognised as a risk factor for delirium (Arizumi et al., 2021). Therefore, these results are not necessarily generalisable to broader postoperative populations outside of critical care environments.

## Conclusion

This study explored the use of ABNs to analyse patient data of the ICU, identify delirium risk factors, and compare these findings to results from statistical analyses and literature. Delirium remains a condition with multifactorial and interdependent relationships among its influencing factors, making prediction and risk assessment particularly challenging.

The study provides a comprehensive overview of risk factors of delirium, the methodological background of ABNs, and demonstrates their application to delirium and related cohorts with similar symptoms. This study also highlights how ABNs can show associations between risk factors and conditions like delirium.

Key findings from the analysis reveal the intricate interplay between mechanical ventilation, age, and antipsychotics with delirium. While some results align with expectations, others, such as the counterintuitive relationship between age, ventilation, and ICDSC ≥4, challenge conventional perspectives. These discrepancies underline the need for further investigation and validation using larger, more comprehensive datasets.

Future research should focus on exploring the complex interactions between ventilation, antipsychotics, and delirium. Expanding datasets and increasing the number of parent variables in ABNs could refine the model and enhance its accuracy. Addressing overfitting through simulations and pruning techniques would further strengthen the reliability of the results. Additionally, exploring the connections between the variables and different types of delirium, such as hypoactive and hyperactive forms, would be interesting, as these types require distinct management strategies.

## Data Availability

All data produced in the present study are available upon reasonable request to the authors

## Acknowledgements

The first author thanks Georg R. Spinner and Jan Bartussek for their guidance and feedback on the project.

## List of Abbreviations

ABN: Additive Bayesian Network
BN: Bayesian Network
CAM-ICU: Confusion AssessmentMethod for the ICU
DAG: Directed Acyclic Graph
DT: Delirium tremens
FiO_2_: Fraction of inspired Oxygen
GLM: Generalised linear model (
HE: Hepatic encephalopathy
ICD: International Statistical Classification of Diseases and Related Health Prob-lems
ICDSC: Intensive Care DeliriumScreening Checklist
ICU: Intensive care unit
MDSI: Minimaler Datensatz der SGI
NRS: Numeric Rating Scale
PaO_2_: Partial pressure of Oxygen
PCT: Procalcitonin
RASS: Richmond Agitation-Sedation Scale
SAPS: Simplified Acute PhysiologyScore
USZ: University Hospital Zurich
VRS: Verbal Rating Scale
ZOPA: Zurich Observation PainAssessment

## Appendix

The appendix includes the following information:

- Blacklist
- Whitelist
- Categorised medication list
- Distribution of data based on ICD Delirium
- Time thresholds of delirium
- ABN maximum parent analysis
- Computation time
- Bayesian Network DAG Plots
- Bayesian Network Coefficients

## Appendix 1 Blacklist

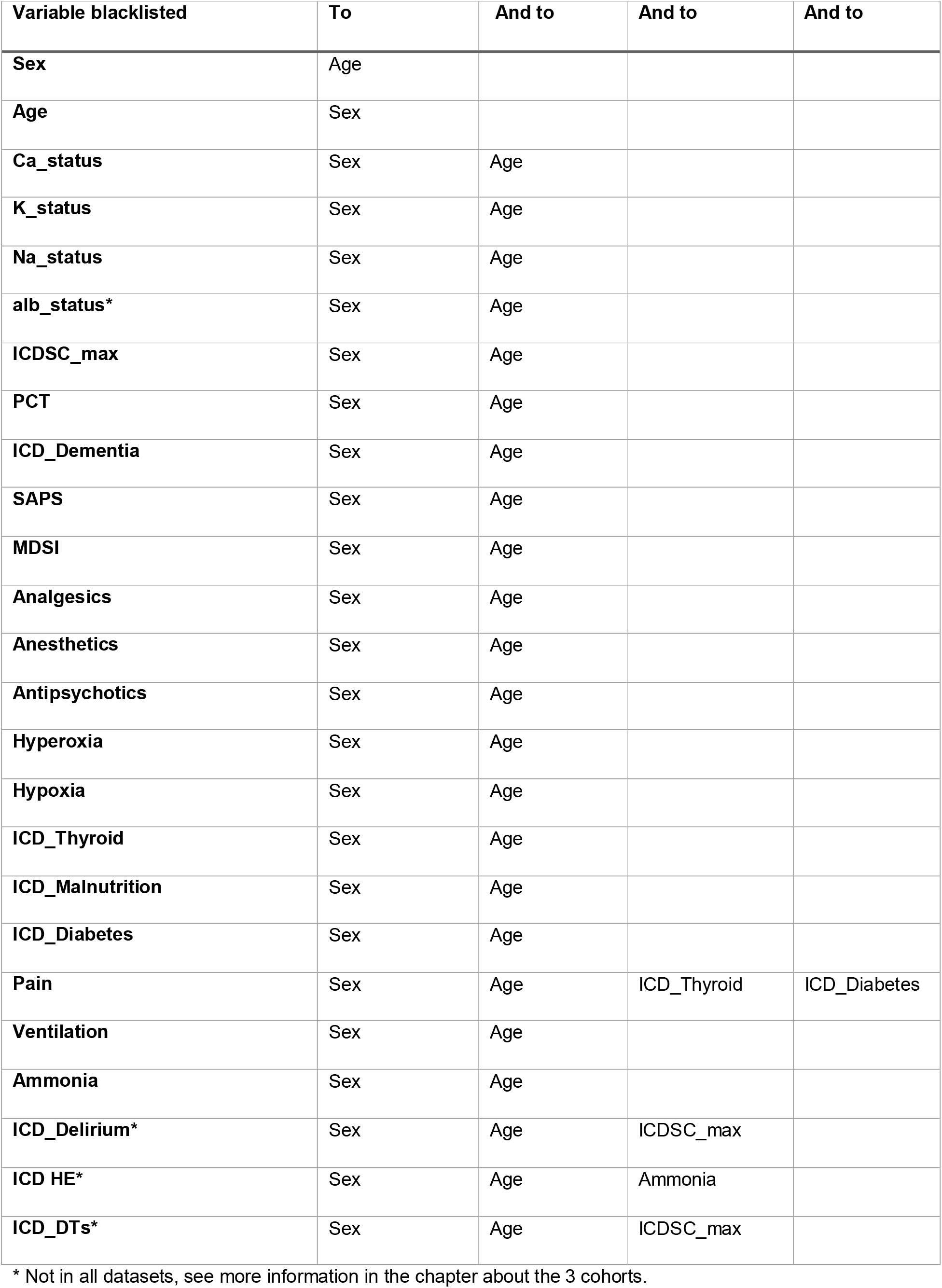

Appendix 1 - Blacklist
| Variable blacklisted | To | And to | And to | And to |
| --- | --- | --- | --- | --- |
| **Sex** | Age |  |  |  |
| **Age** | Sex |  |  |  |
| **Ca_status** | Sex | Age |  |  |
| **K_status** | Sex | Age |  |  |
| **Na_status** | Sex | Age |  |  |
| **alb_status\*** | Sex | Age |  |  |
| **ICDSC_max** | Sex | Age |  |  |
| **PCT** | Sex | Age |  |  |
| **ICD_Dementia** | Sex | Age |  |  |
| **SAPS** | Sex | Age |  |  |
| **MDSI** | Sex | Age |  |  |
| **Analgesics** | Sex | Age |  |  |
| **Anesthetics** | Sex | Age |  |  |
| **Antipsychotics** | Sex | Age |  |  |
| **Hyperoxia** | Sex | Age |  |  |
| **Hypoxia** | Sex | Age |  |  |
| **ICD_Thyroid** | Sex | Age |  |  |
| **ICD_Malnutrition** | Sex | Age |  |  |
| **ICD_Diabetes** | Sex | Age |  |  |
| **Pain** | Sex | Age | ICD_Thyroid | ICD_Diabetes |
| **Ventilation** | Sex | Age |  |  |
| **Ammonia** | Sex | Age |  |  |
| **ICD_Delirium\*** | Sex | Age | ICDSC_max |  |
| **ICD HE\*** | Sex | Age | Ammonia |  |
| **ICD_DTs\*** | Sex | Age | ICDSC_max |  |

\* Not in all datasets, see more information in the chapter about the 3 cohorts.

## Appendix 2 Whitelist

Appendix 2 - Whitelist
| from | to |
| --- | --- |
| Age | SAPS |
| Na_Status | SAPS |
| K_Status | SAPS |

## Appendix 3 Categorised medication list

Appendix 3 – Categorised medication list
| Medication | Category |
| --- | --- |
| Morphin.sulfat.pentahydrat..mg. | Analgesics |
| Morphin.hydrochlorid.3.Wasser..mg. | Analgesics |
| Morphin.hydrochlorid..mg. | Analgesics |
| Fentanyl..mcg. | Analgesics |
| Codein.phosphat.hemihydrat..mg. | Analgesics |
| Clonidin.hydrochlorid..mg. | Analgesics |
| Amitriptylin..mg. | Analgesics |
| Thiopental.natrium..mg. | Anesthetics |
| Sufentanil..mg. | Anesthetics |
| Ropivacain.hydrochlorid..500mg....Fentanyl..500mcg. | Anesthetics |
| Propofol..mg. | Anesthetics |
| Midazolam..mg. | Anesthetics |
| Ketamin..mg. | Anesthetics |
| Dexmedetomidin..mg. | Anesthetics |
| Risperidon..mg. | Antipsychotics |
| Quetiapin..mg. | Antipsychotics |
| Olanzapin..mg. | Antipsychotics |
| Haloperidol..mg. | Antipsychotics |

## Appendix 4 - Distribution of data based on ICD Delirium

|  | No Delirium (n=864) | Delirium (n=514) |
| --- | --- | --- |
| Age, median, yr | 63 years (21-97) | 70 years (20-100) |
| Sex, M, n (%) | 528 (61%) | 376 (73%) |
| Calcium | both: 7, hyper: 37, hypo: 491, normal: 329 | both: 6, hyper: 32, hypo: 276, normal: 200 |
| Potassium | both: 13, hyper: 206, hypo: 178, normal: 467 | both: 7, hyper: 146, hypo: 82, normal: 279 |
| Sodium | both: 3, hyper: 316, hypo: 92, normal: 453 | both: 2, hyper: 222, hypo: 39, normal: 251 |
| **Albumin** | hypo: 847, normal: 17 | hypo: 510, normal: 4 |
| **PCT, 1, n (%)** | 744 (86%) | 409 (80%) |
| **ICD Dementia, 1, n (%)** | 9 (1%) | 15 (3%) |
| **SAPS, median** | 47 (2-126) | 50.5 (0-101) |
| **Mechanically ventilated during ICU stay, 1, n (%)** | 586 (67%) | 341 (66%) |
| **MDSI** | 1: 239<br>2: 161<br>3: 90<br>4: 217<br>5: 17<br>6: 62<br>7: 3<br>8: 75 | 1: 199<br>2: 76<br>3: 46<br>4: 85<br>5: 7<br>6: 55<br>7: 7<br>8: 39 |
| **Analgesics 1, n (%)** | 451 (52%) | 277 (54%) |
| **Anaesthetics, 1, n (%)** | 508 (59%) | 359 (70%) |
| **Antipsychotics, 1, n (%)** | 89 (10%) | 239 (46%) |
| **Hyperoxia** | None: 449<br>mild: 220<br>moderate: 116<br>severe: 79 | None: 262<br>mild: 143<br>moderate: 70<br>severe: 39 |
| **Hypoxia** | mild: 368, moderate: 86, None: 352, severe: 58 | mild: 234, moderate: 60, None: 171, severe: 49 |
| **ICD Thyroid, 1, n (%)** | 57 (7%) | 45 (9%) |
| **ICD Malnutrition, 1, n (%)** | 75 (9%) | 61 (12%) |
| **ICD Diabetes, 1, n (%)** | 155 (18%) | 142 (28%) |
| **Pain, 1, n (%)** | 315 (36%) | 179 (35%) |

## Appendix 5 ICDSC Time Thresholds

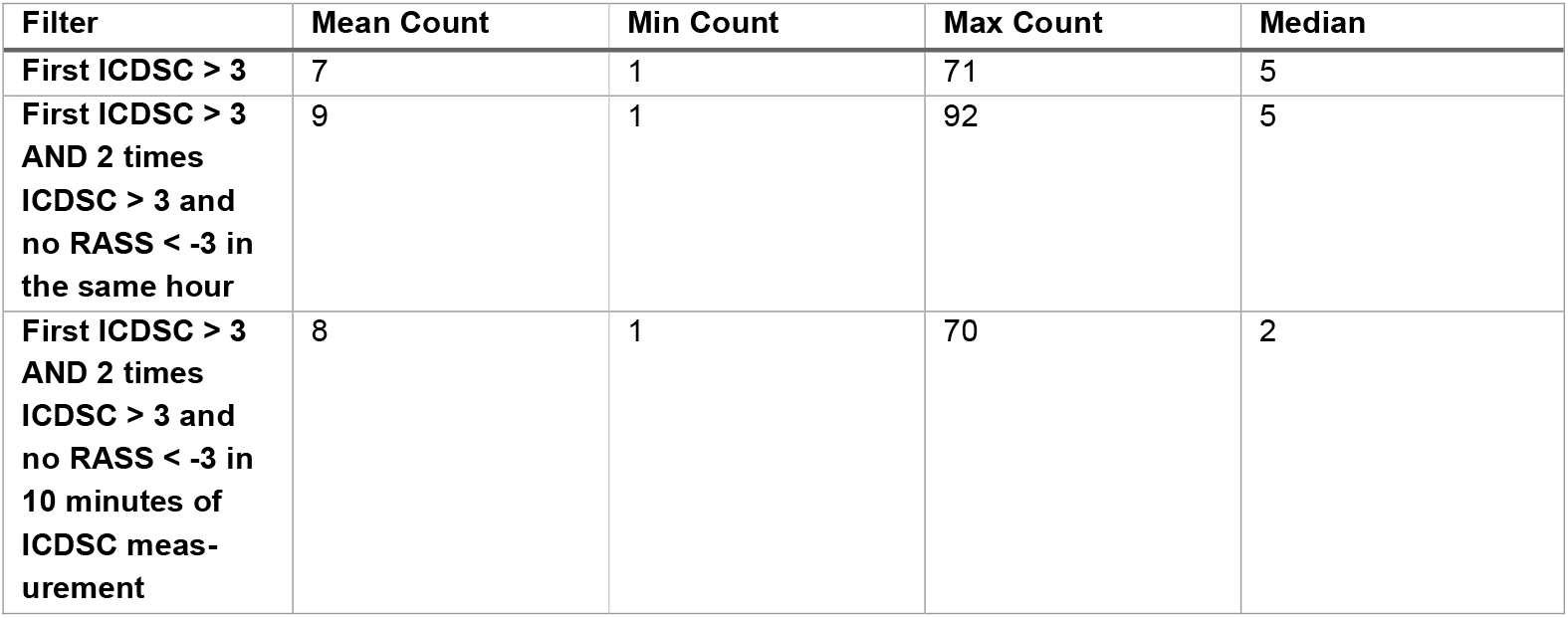

## Appendix 6 ABN maximum parent analysis

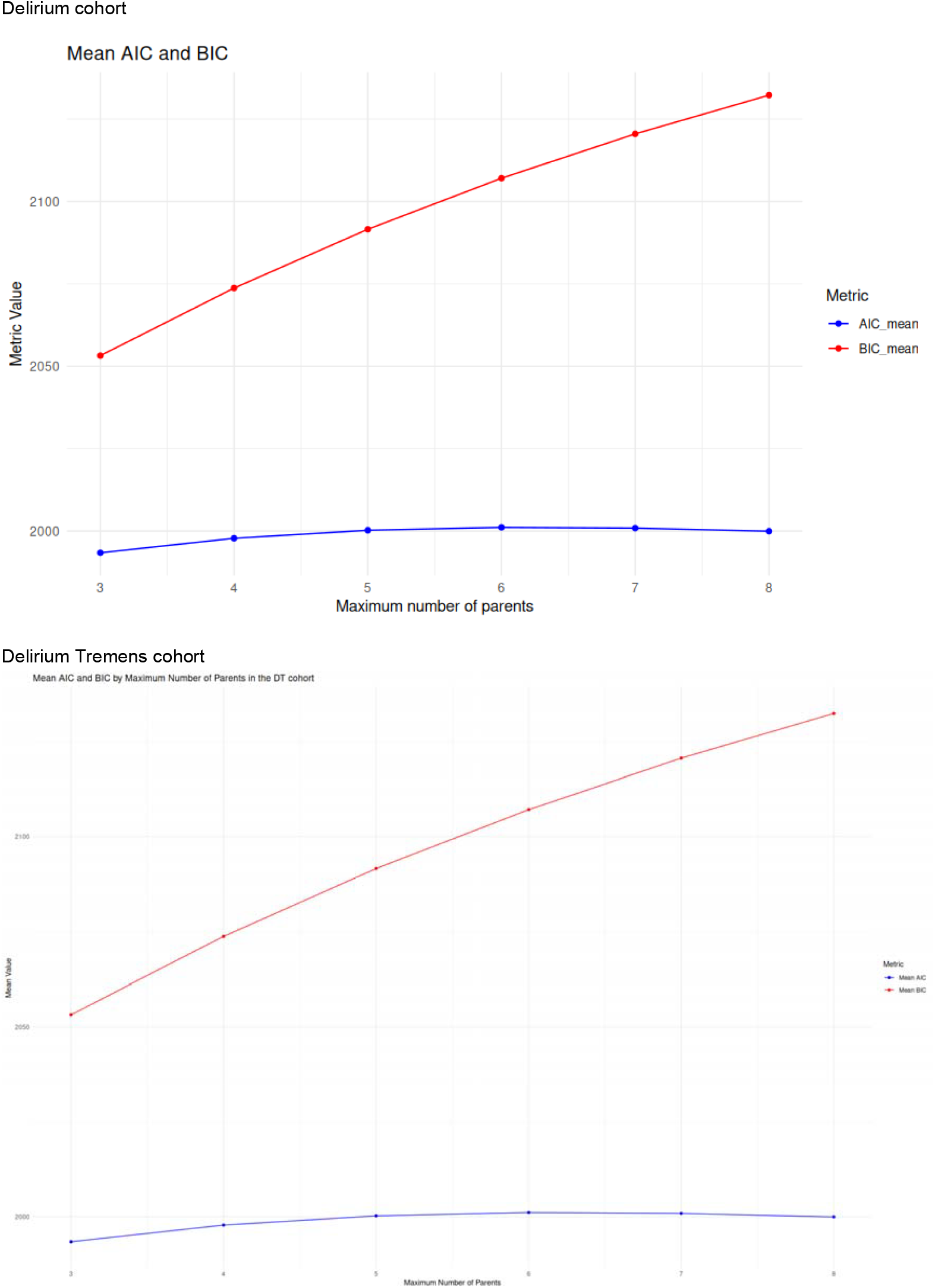
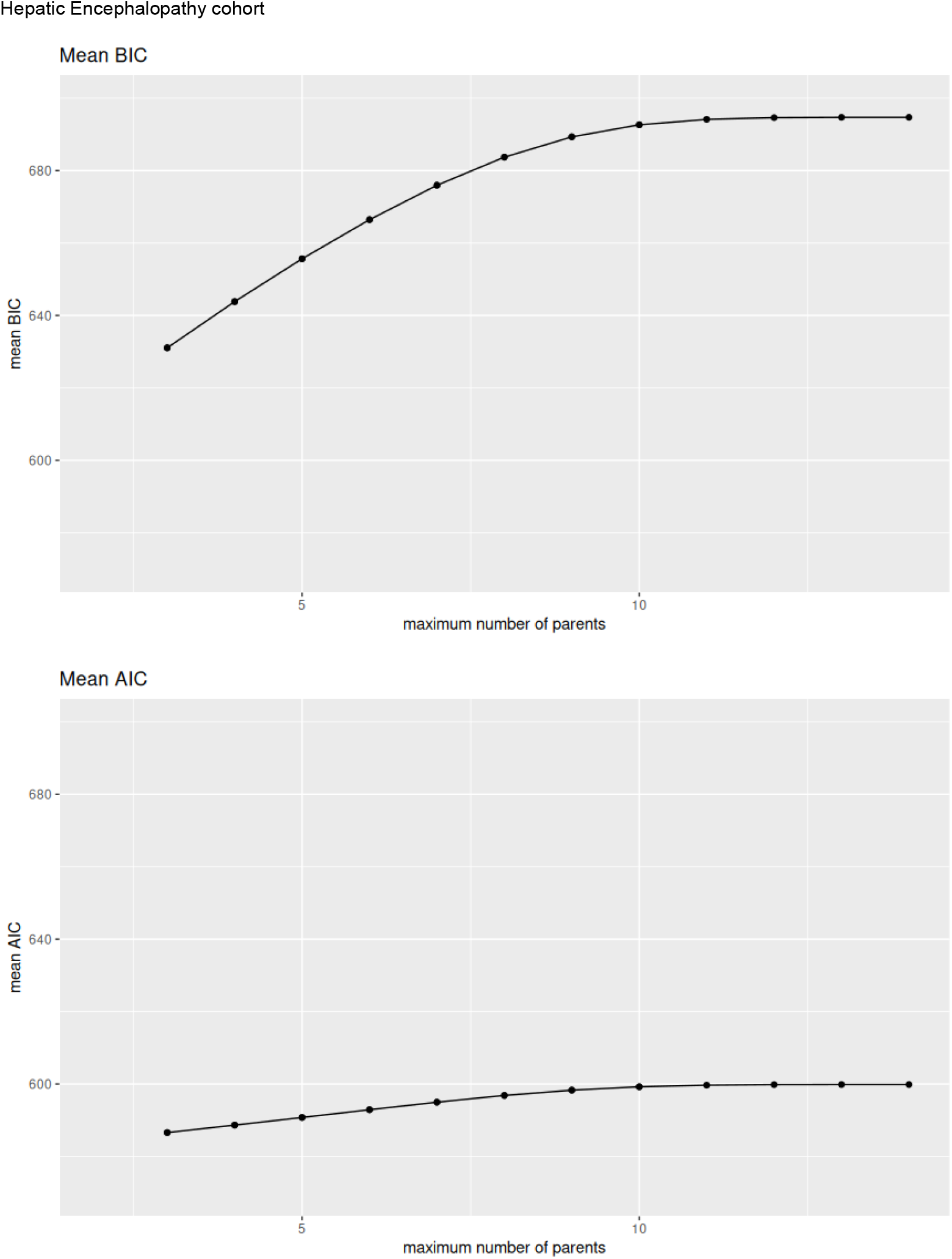

## Appendix 7 Computation time

Appendix 7 – Computation time
| Cohort | Max Parent | Calculation | Cores | Time approximately |
| --- | --- | --- | --- | --- |
| **Delirium Tremens 22 parameter, 900 patients** | 4 | Abn Score Cache | 3 | <10 min |
| **Delirium Tremens 22 parameter, 900 patients** | 4 | DAG | 1 | 25 hours |
| **Delirium, 22 Parameters, 1200 patients** | 4 | Abn Score Cache | 3 | <10 min |
| **Delirium, 22 Parameters, 1200 patients** | 4 | DAG | 1 | 25 hours |
| **Delirium Tremens 22 parameter, 900 patients** | 6 | Abn Score Cache | 3 | 2 hours |
| **Delirium Tremens 22 parameter, 900 patients** | 6 | DAG | 1 | 40 hours |
| **Delirium Tremens 22 parameter, 900 patients** | 7 | Abn Score Cache | 3 | 5 hours |
| **Delirium Tremens 22 parameter, 900 patients** | 7 | DAG | 1 | 3 days |
| **Delirium, 22 Parameters, 1200 patients** | 7 | Abn Score Cache | 3 | 10 hours |
| **Delirium, 22 Parameters, 1200 patients** | 7 | DAG | 1 | 3 days |
| **Delirium, 22 Parameters, 1200 patients** | 8 | Abn Score Cache | 3 | 20 hours |
| **Delirium, 22 Parameters, 1200 patients** | 8 | DAG | 1 | 6 Tage |
| **HE, 16 parameters, 200 patients** | 15 | Abn Score Cache | 3 | 15 min |
| **HE, 16 parameters, 200 patients** | 15 | DAG | 1 | 5 min |

## Appendix 8 Bayesian Network DAG plots

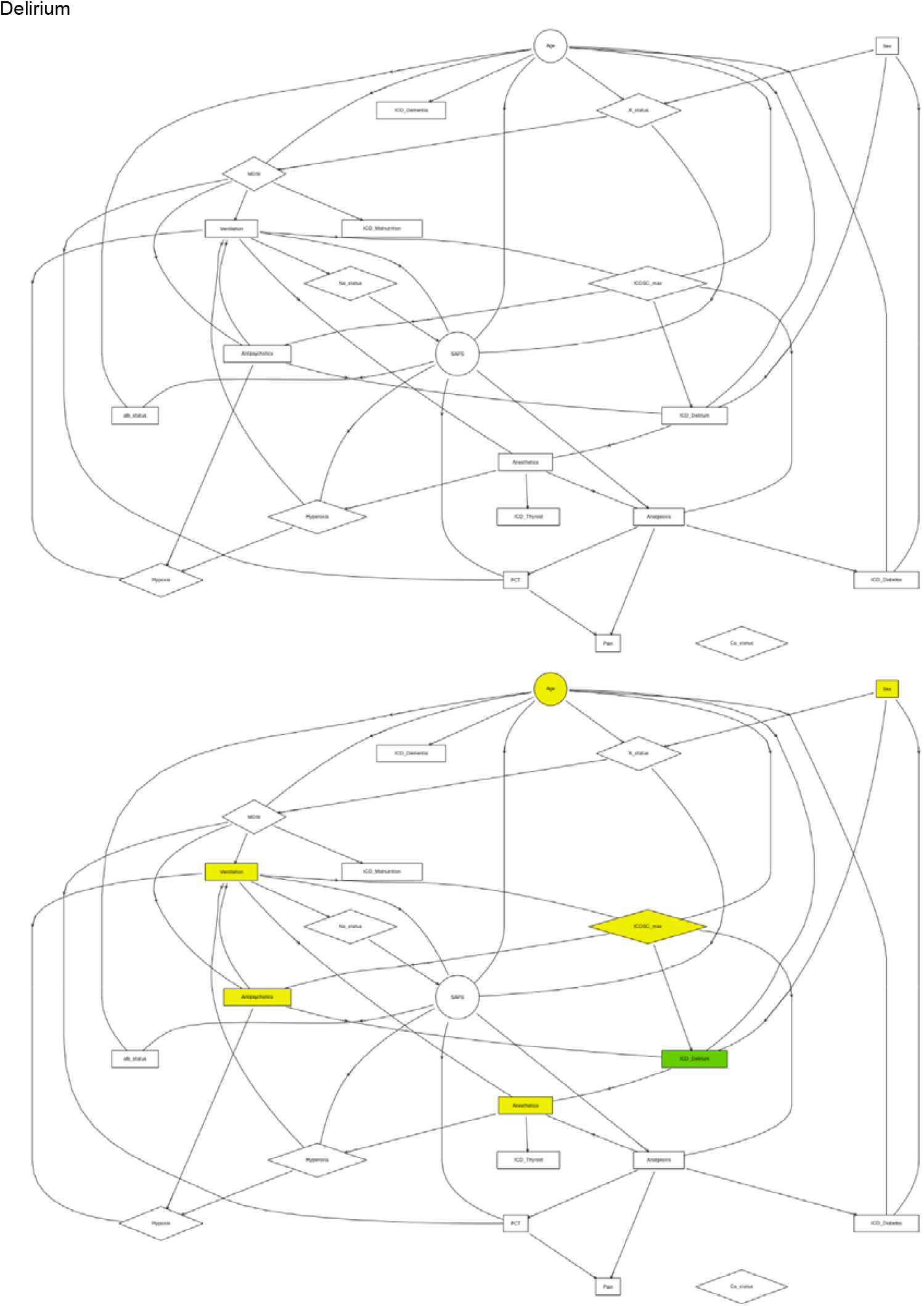
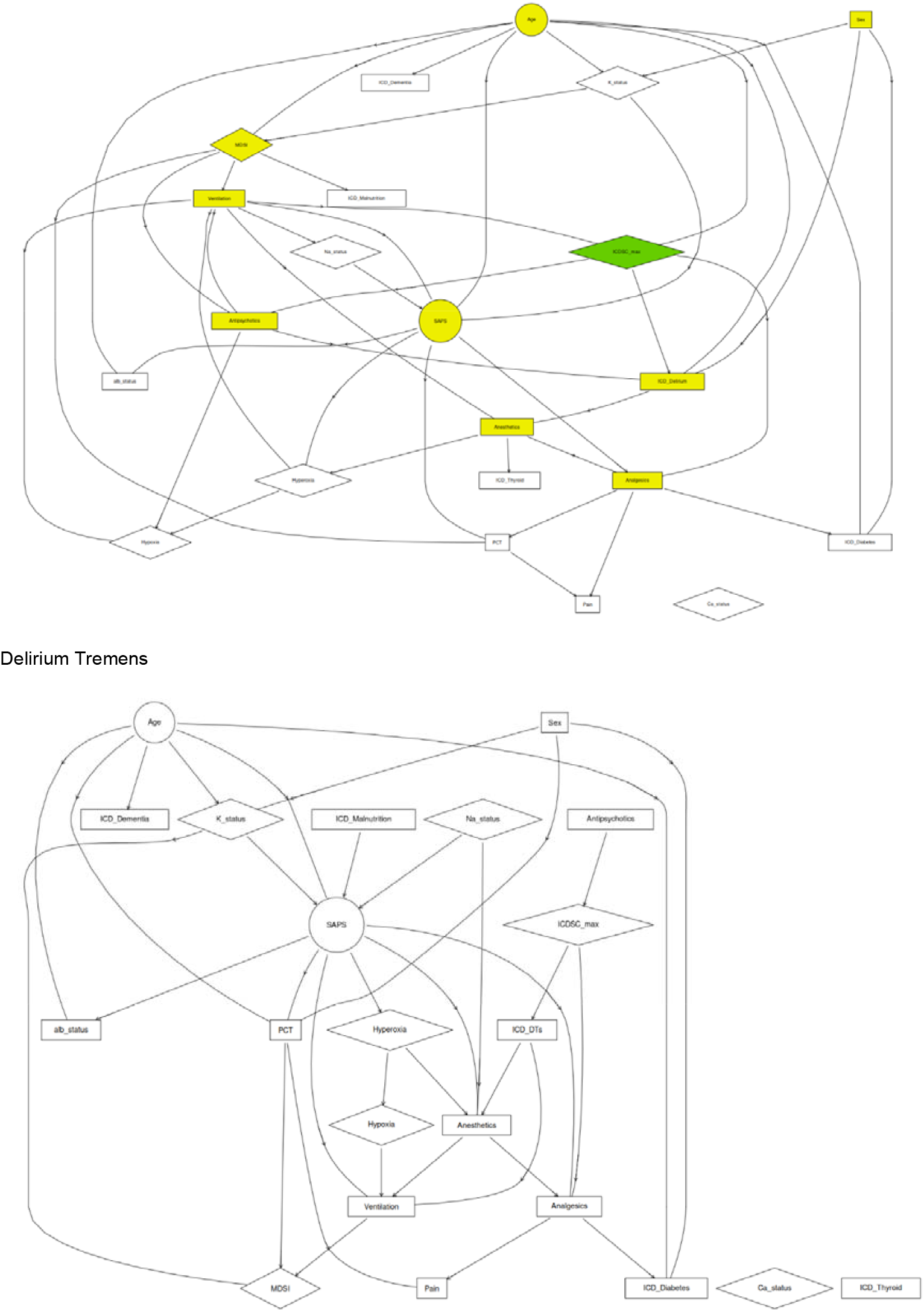
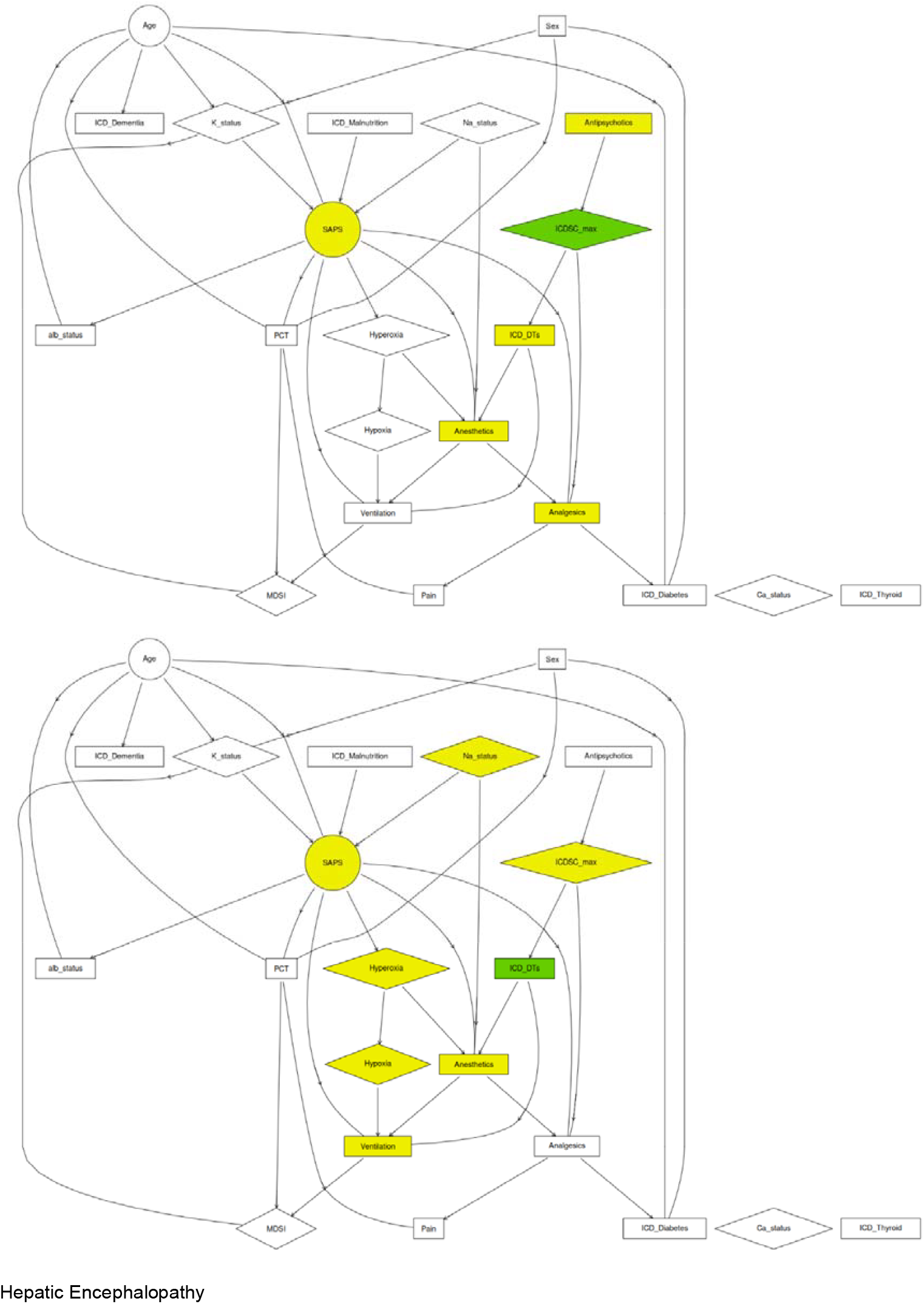
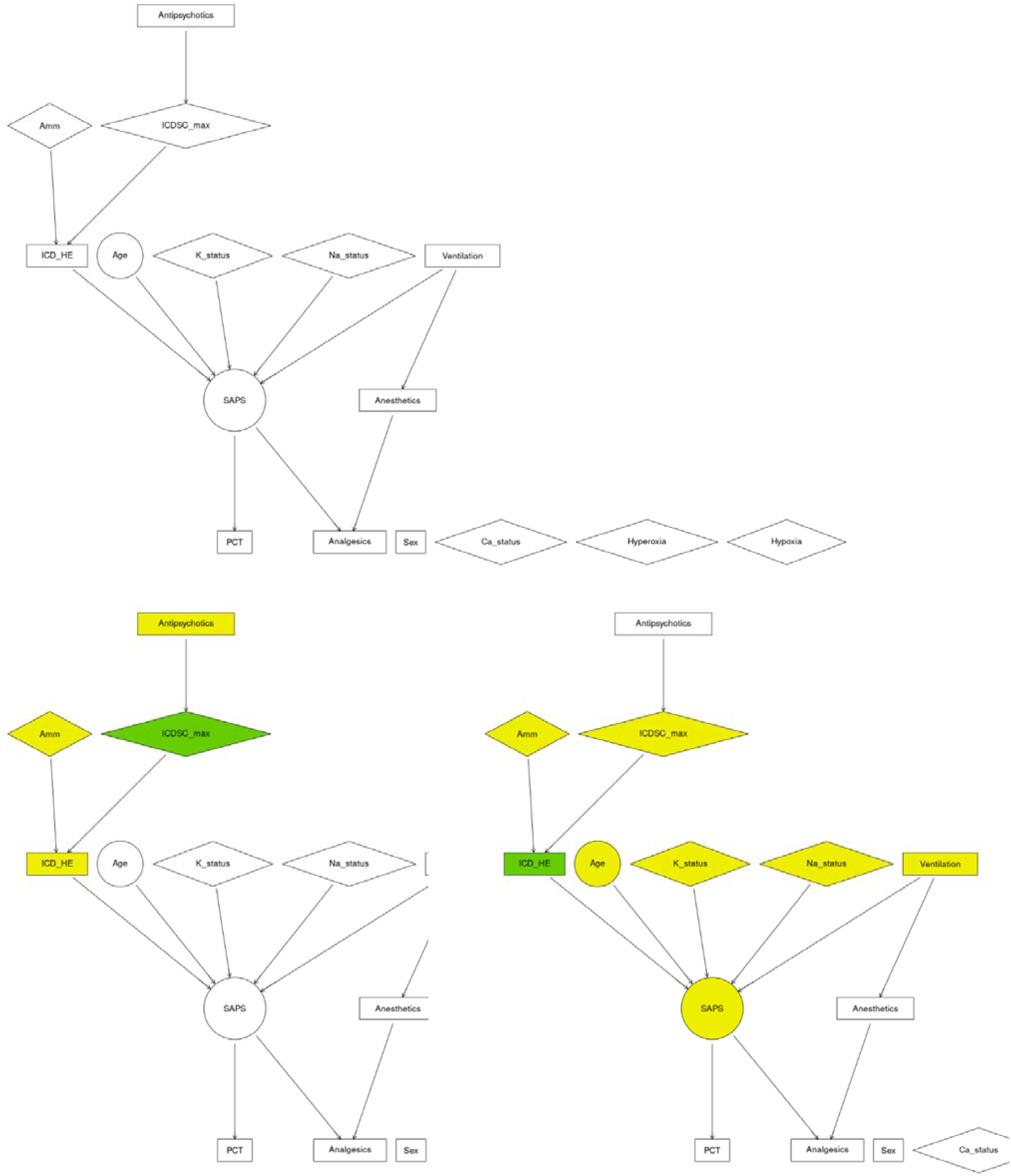

## Appendix 9 Bayesian Network coefficients

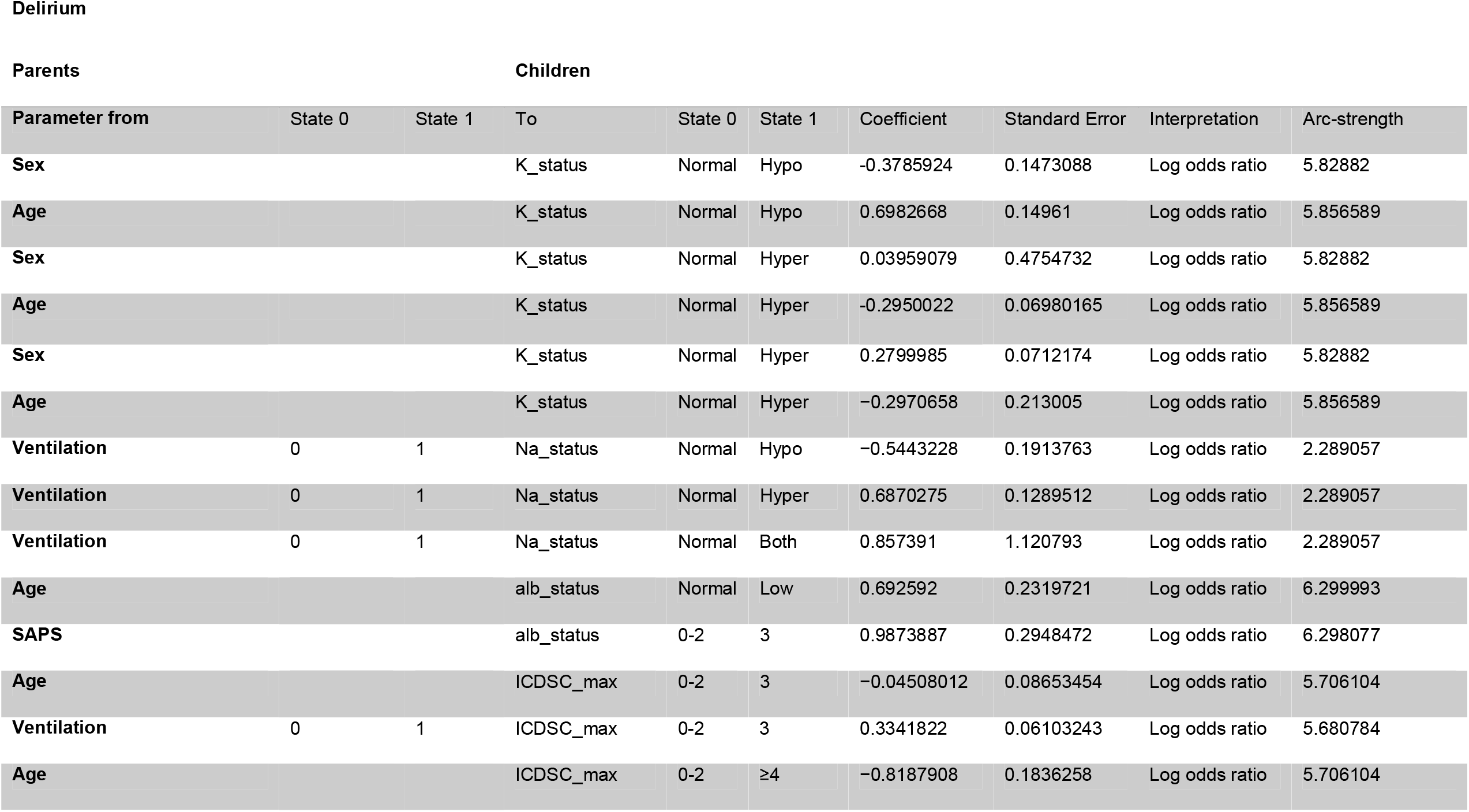
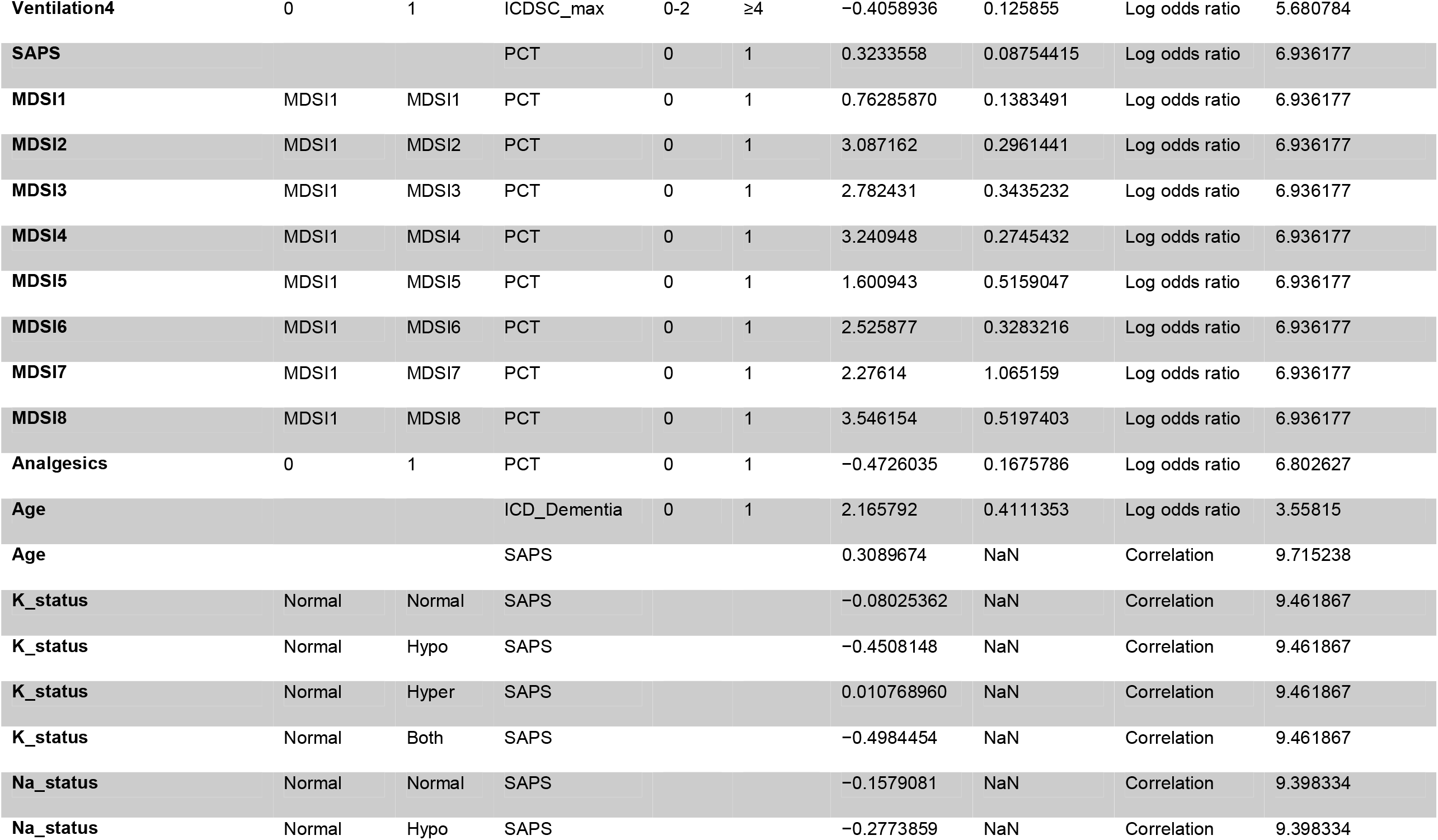
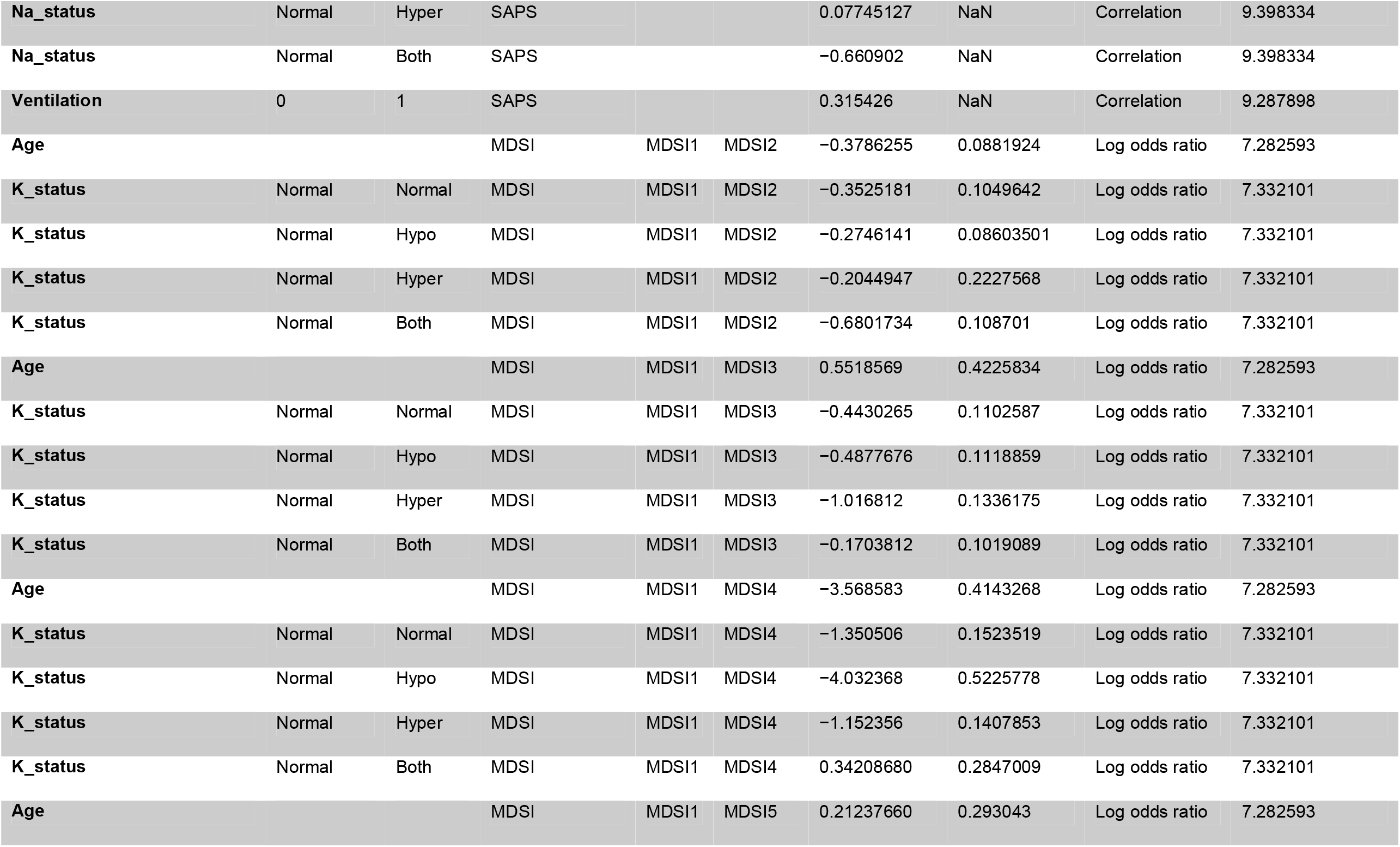
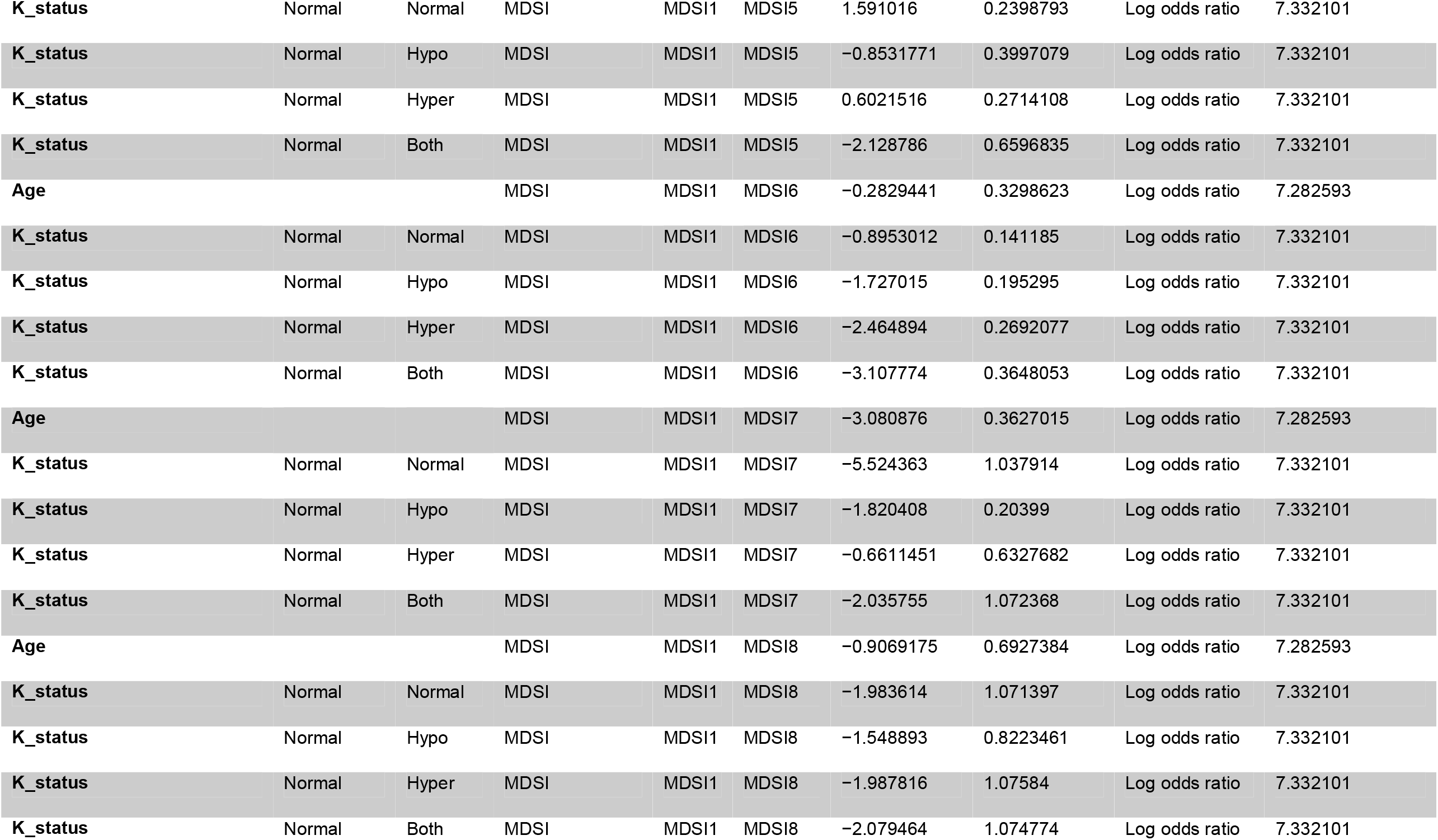
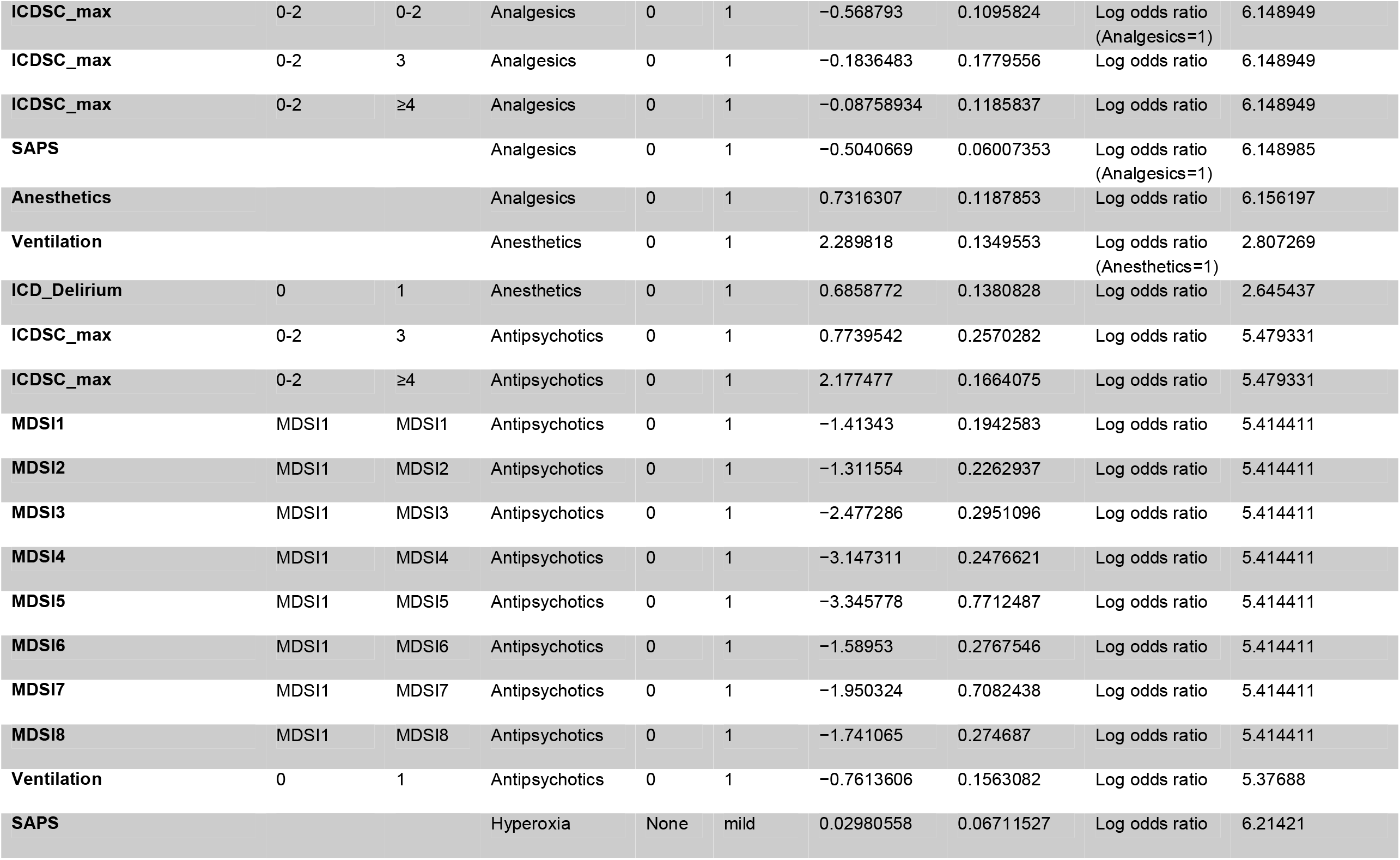
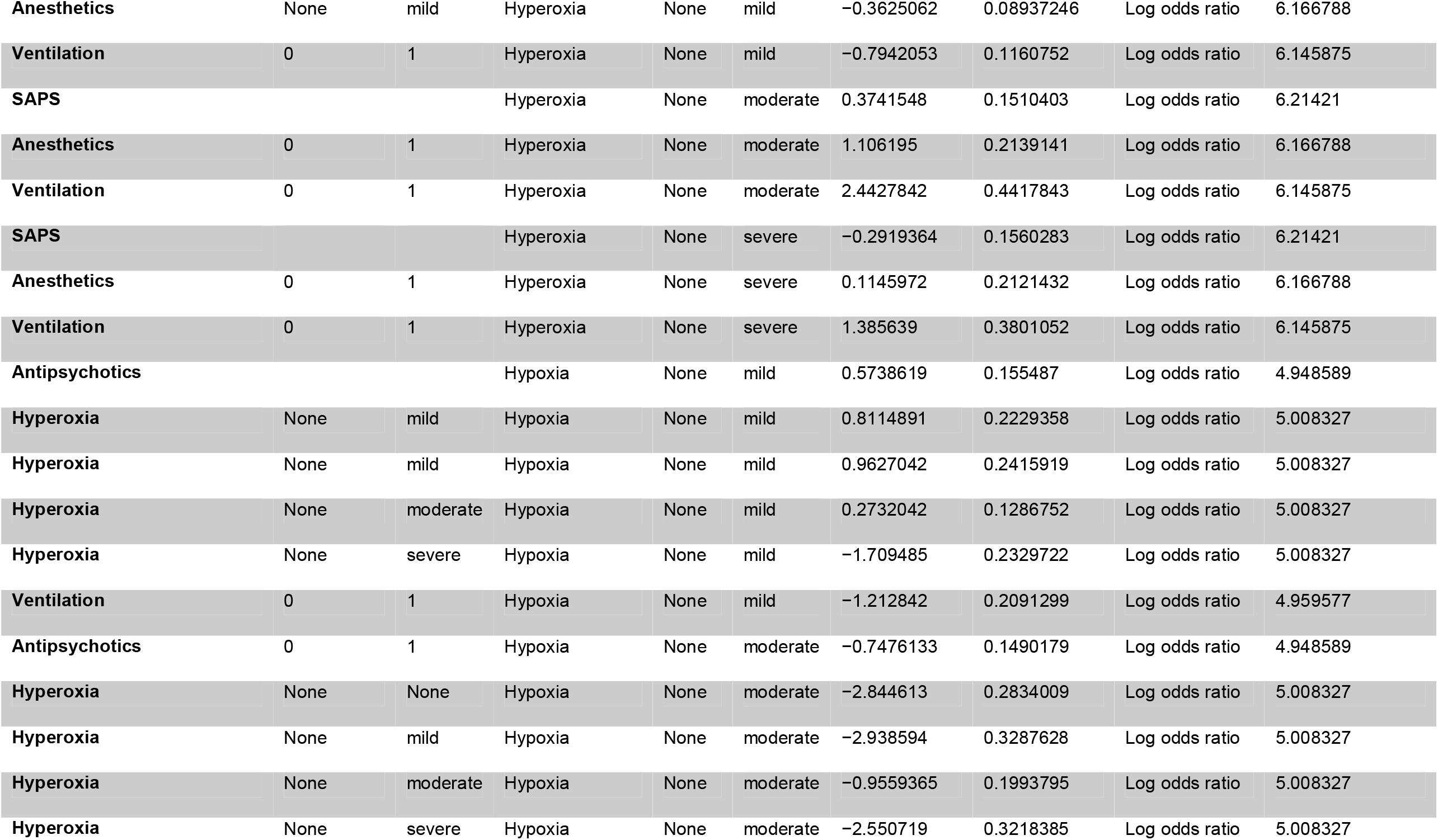
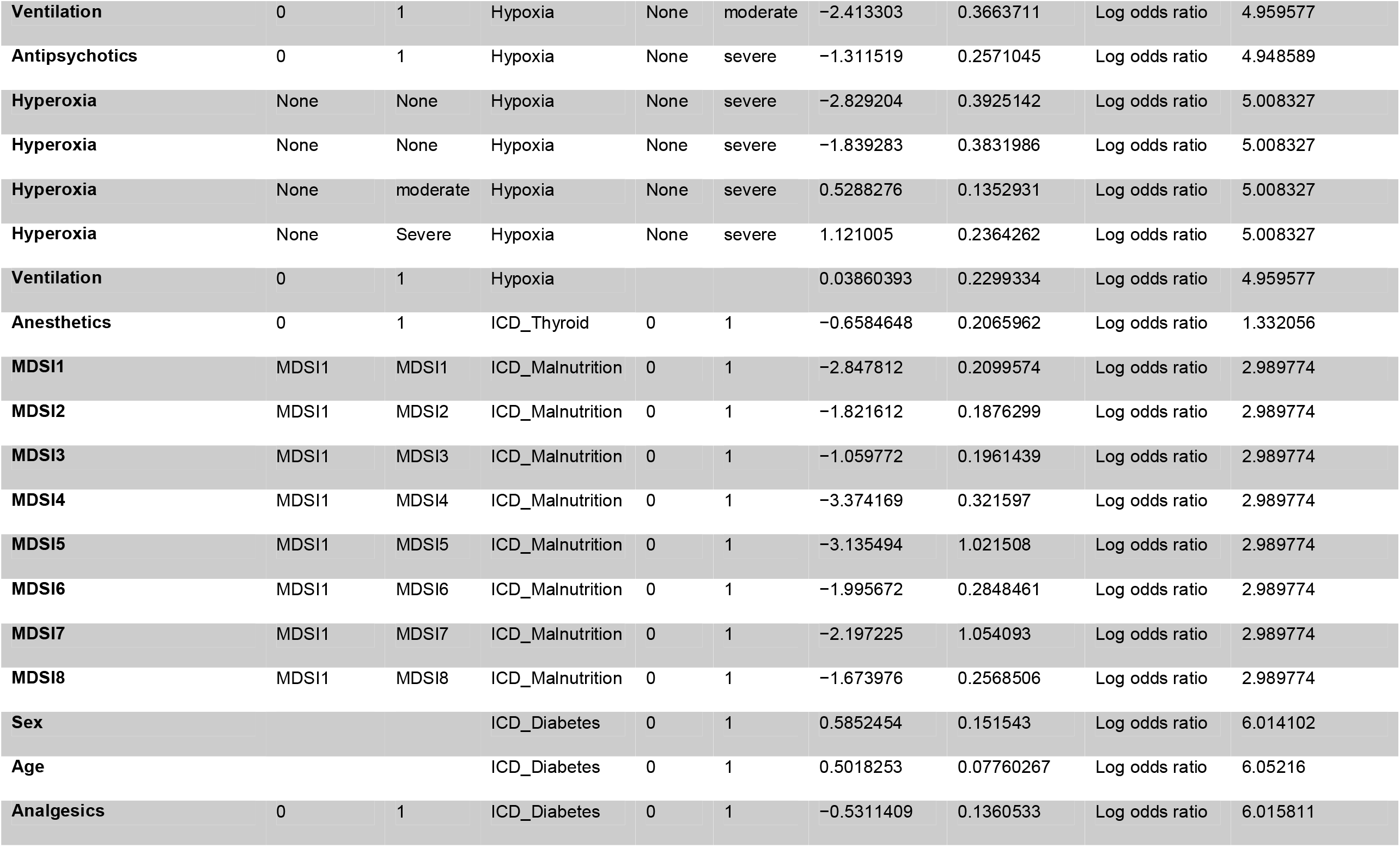
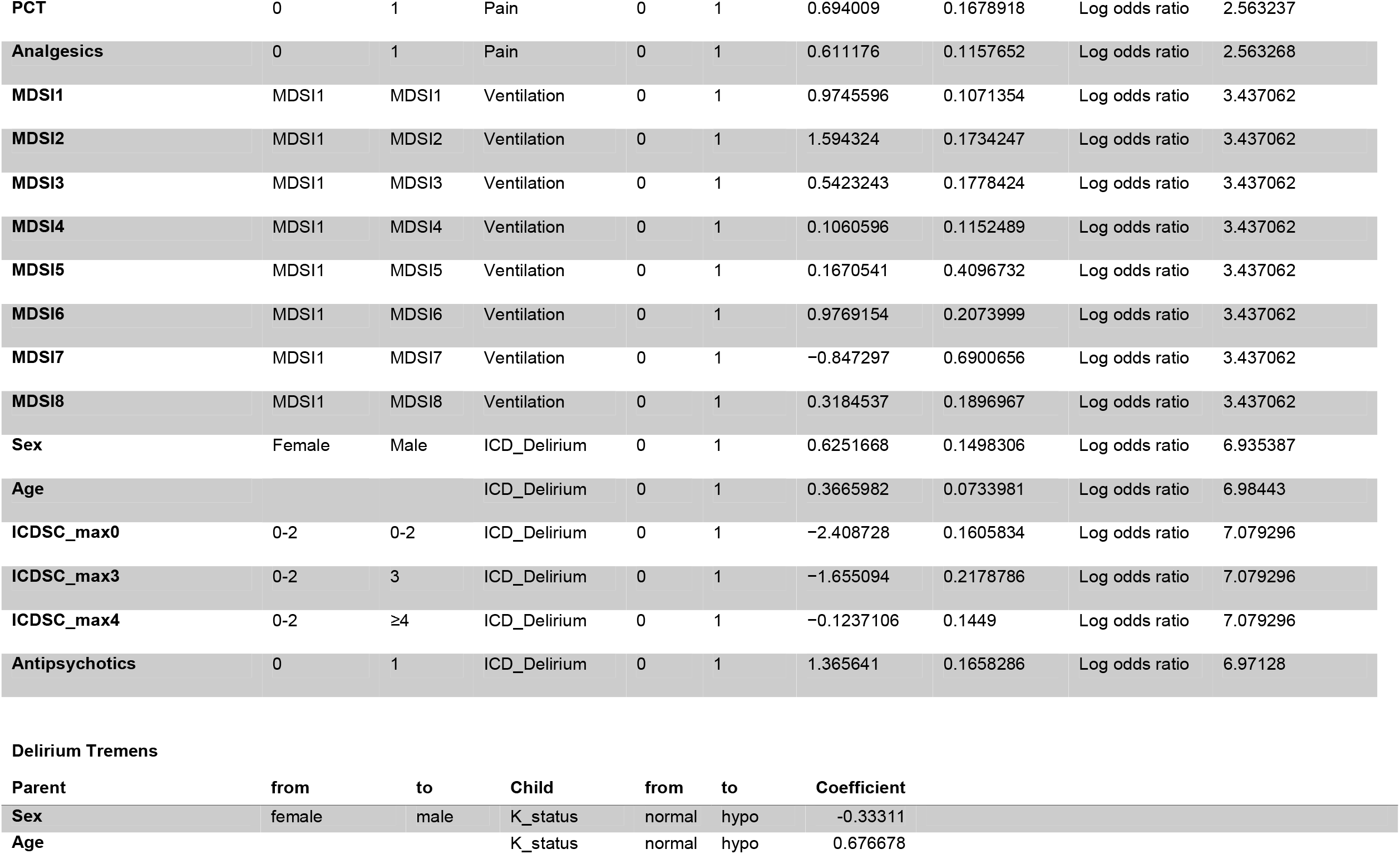
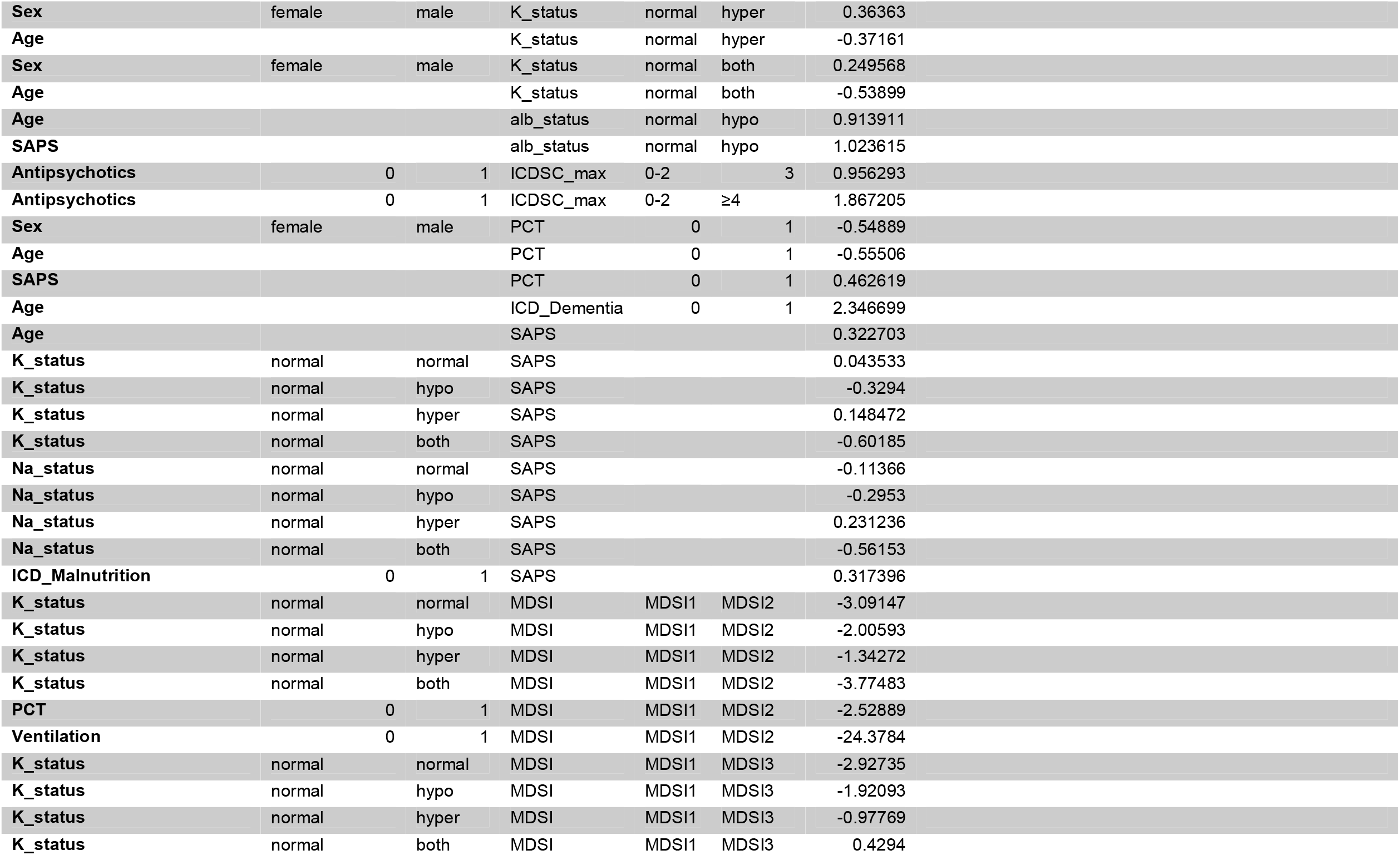
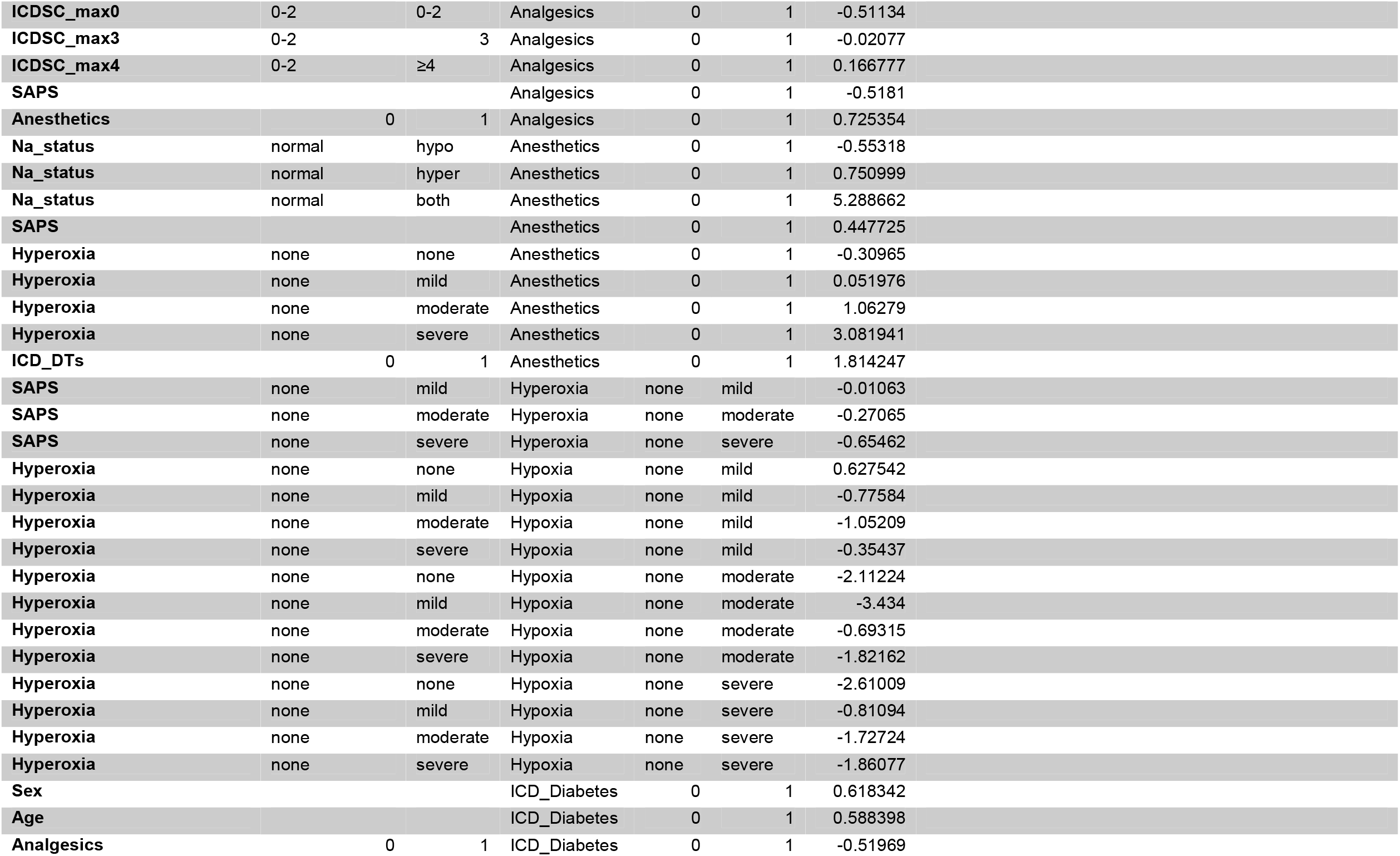
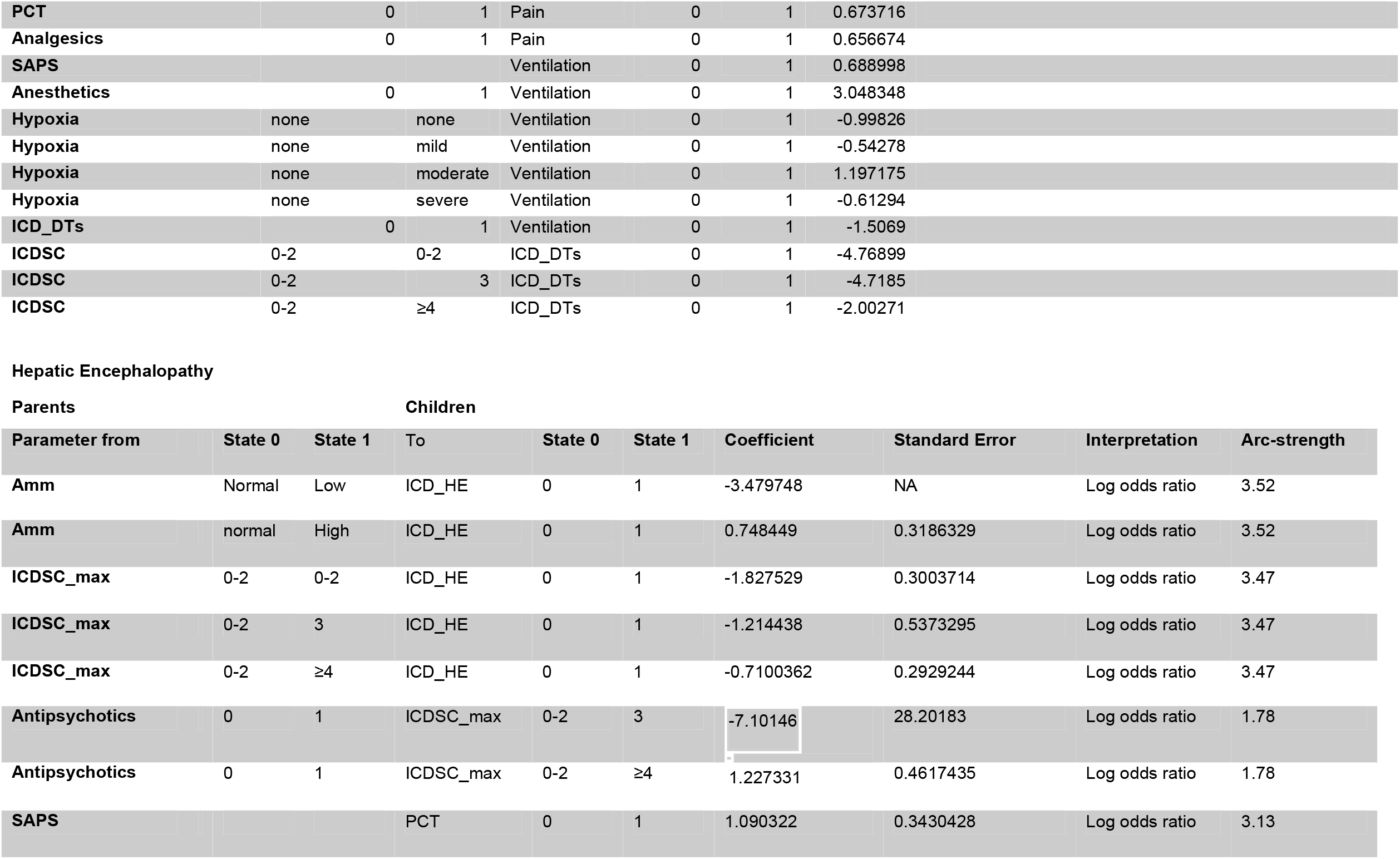
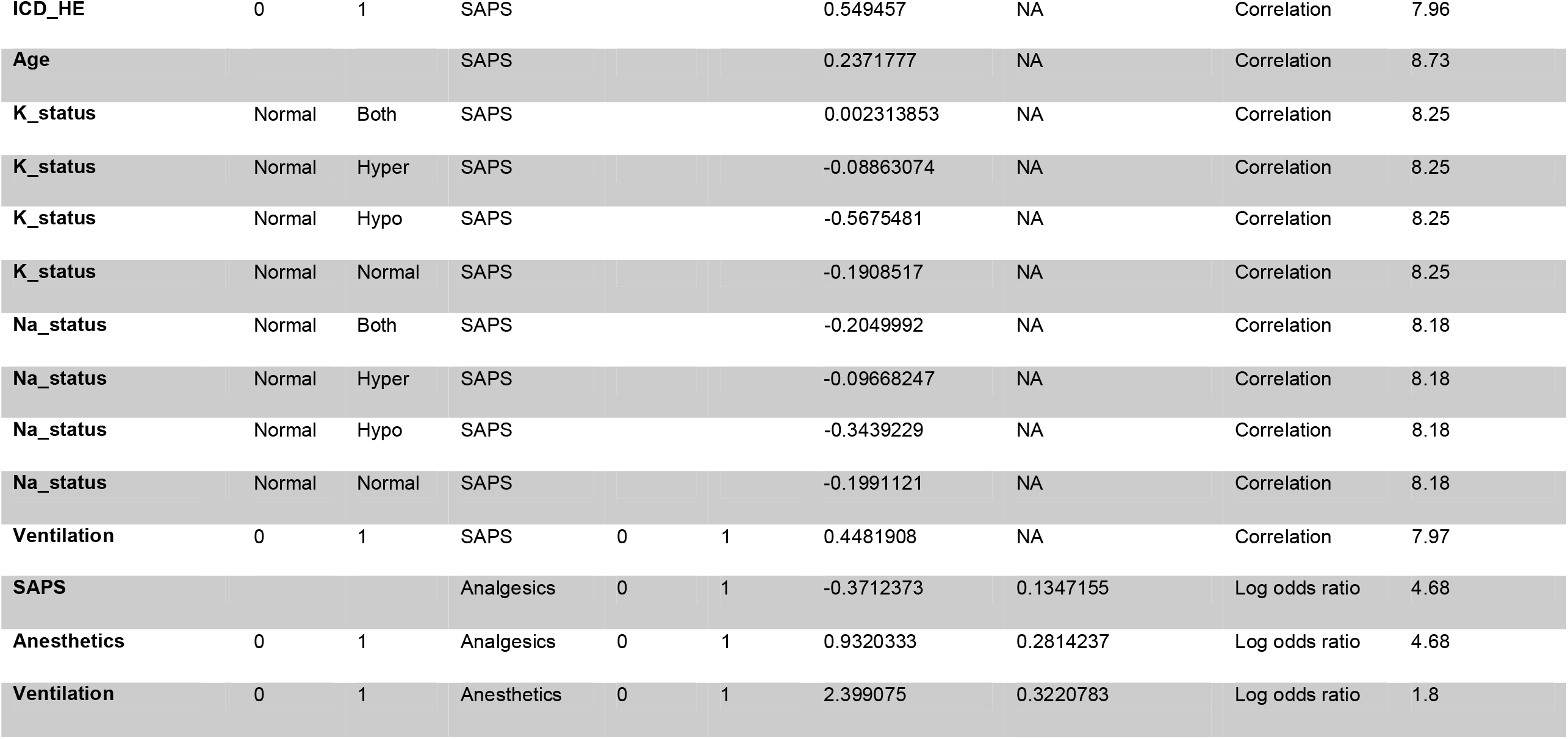

Children
| Parameter from | State 0 | State 1 | To | State 0 | State 1 | Coefficient | Standard Error | Interpretation | Arc-strength |
| --- | --- | --- | --- | --- | --- | --- | --- | --- | --- |
| Sex |  |  | K_status | Normal | Hypo | -0.3785924 | 0.1473088 | Log odds ratio | 5.82882 |
| Age |  |  | K_status | Normal | Hypo | 0.6982668 | 0.14961 | Log odds ratio | 5.856589 |
| Sex |  |  | K_status | Normal | Hyper | 0.03959079 | 0.4754732 | Log odds ratio | 5.82882 |
| Age |  |  | K_status | Normal | Hyper | -0.2950022 | 0.06980165 | Log odds ratio | 5.856589 |
| Sex |  |  | K_status | Normal | Hyper | 0.2799985 | 0.0712174 | Log odds ratio | 5.82882 |
| Age |  |  | K_status | Normal | Hyper | -0.2970658 | 0.213005 | Log odds ratio | 5.856589 |
| Ventilation | 0 | 1 | Na_status | Normal | Hypo | -0.5443228 | 0.1913763 | Log odds ratio | 2.289057 |
| Ventilation | 0 | 1 | Na_status | Normal | Hyper | 0.6870275 | 0.1289512 | Log odds ratio | 2.289057 |
| Ventilation | 0 | 1 | Na_status | Normal | Both | 0.857391 | 1.120793 | Log odds ratio | 2.289057 |
| Age |  |  | alb_status | Normal | Low | 0.692592 | 0.2319721 | Log odds ratio | 6.299993 |
| SAPS |  |  | alb_status | 0-2 | 3 | 0.9873887 | 0.2948472 | Log odds ratio | 6.298077 |
| Age |  |  | ICDSC_max | 0-2 | 3 | -0.04508012 | 0.08653454 | Log odds ratio | 5.706104 |
| Ventilation | 0 | 1 | ICDSC_max | 0-2 | 3 | 0.3341822 | 0.06103243 | Log odds ratio | 5.680784 |
| Age |  |  | ICDSC_max | 0-2 | ≥4 | -0.8187908 | 0.1836258 | Log odds ratio | 5.706104 |
| **Ventilation4** | 0 | 1 | ICDSC_max | 0-2 | ≥4 | -0.4058936 | 0.125855 | Log odds ratio | 5.680784 |
| **SAPS** |  |  | PCT | 0 | 1 | 0.3233558 | 0.08754415 | Log odds ratio | 6.936177 |
| **MDSI1** | MDSI1 | MDSI1 | PCT | 0 | 1 | 0.76285870 | 0.1383491 | Log odds ratio | 6.936177 |
| **MDSI2** | MDSI1 | MDSI2 | PCT | 0 | 1 | 3.087162 | 0.2961441 | Log odds ratio | 6.936177 |
| **MDSI3** | MDSI1 | MDSI3 | PCT | 0 | 1 | 2.782431 | 0.3435232 | Log odds ratio | 6.936177 |
| **MDSI4** | MDSI1 | MDSI4 | PCT | 0 | 1 | 3.240948 | 0.2745432 | Log odds ratio | 6.936177 |
| **MDSI5** | MDSI1 | MDSI5 | PCT | 0 | 1 | 1.600943 | 0.5159047 | Log odds ratio | 6.936177 |
| **MDSI6** | MDSI1 | MDSI6 | PCT | 0 | 1 | 2.525877 | 0.3283216 | Log odds ratio | 6.936177 |
| **MDSI7** | MDSI1 | MDSI7 | PCT | 0 | 1 | 2.27614 | 1.065159 | Log odds ratio | 6.936177 |
| **MDSI8** | MDSI1 | MDSI8 | PCT | 0 | 1 | 3.546154 | 0.5197403 | Log odds ratio | 6.936177 |
| **Analgesics** | 0 | 1 | PCT | 0 | 1 | -0.4726035 | 0.1675786 | Log odds ratio | 6.802627 |
| **Age** |  |  | ICD_Dementia | 0 | 1 | 2.165792 | 0.4111353 | Log odds ratio | 3.55815 |
| **Age** |  |  | SAPS |  |  | 0.3089674 | NaN | Correlation | 9.715238 |
| **K_status** | Normal | Normal | SAPS |  |  | -0.08025362 | NaN | Correlation | 9.461867 |
| **K_status** | Normal | Hypo | SAPS |  |  | -0.4508148 | NaN | Correlation | 9.461867 |
| **K_status** | Normal | Hyper | SAPS |  |  | 0.010768960 | NaN | Correlation | 9.461867 |
| **K_status** | Normal | Both | SAPS |  |  | -0.4984454 | NaN | Correlation | 9.461867 |
| **Na_status** | Normal | Normal | SAPS |  |  | -0.1579081 | NaN | Correlation | 9.398334 |
| **Na_status** | Normal | Hypo | SAPS |  |  | -0.2773859 | NaN | Correlation | 9.398334 |
| **Na_status** | Normal | Hyper | SAPS |  |  | 0.07745127 | NaN | Correlation | 9.398334 |
| **Na_status** | Normal | Both | SAPS |  |  | -0.660902 | NaN | Correlation | 9.398334 |
| **Ventilation** | 0 | 1 | SAPS |  |  | 0.315426 | NaN | Correlation | 9.287898 |
| **Age** |  |  | MDSI | MDSI1 | MDSI2 | -0.3786255 | 0.0881924 | Log odds ratio | 7.282593 |
| **K_status** | Normal | Normal | MDSI | MDSI1 | MDSI2 | -0.3525181 | 0.1049642 | Log odds ratio | 7.332101 |
| **K_status** | Normal | Hypo | MDSI | MDSI1 | MDSI2 | -0.2746141 | 0.08603501 | Log odds ratio | 7.332101 |
| **K_status** | Normal | Hyper | MDSI | MDSI1 | MDSI2 | -0.2044947 | 0.2227568 | Log odds ratio | 7.332101 |
| **K_status** | Normal | Both | MDSI | MDSI1 | MDSI2 | -0.6801734 | 0.108701 | Log odds ratio | 7.332101 |
| **Age** |  |  | MDSI | MDSI1 | MDSI3 | 0.5518569 | 0.4225834 | Log odds ratio | 7.282593 |
| **K_status** | Normal | Normal | MDSI | MDSI1 | MDSI3 | -0.4430265 | 0.1102587 | Log odds ratio | 7.332101 |
| **K_status** | Normal | Hypo | MDSI | MDSI1 | MDSI3 | -0.4877676 | 0.1118859 | Log odds ratio | 7.332101 |
| **K_status** | Normal | Hyper | MDSI | MDSI1 | MDSI3 | -1.016812 | 0.1336175 | Log odds ratio | 7.332101 |
| **K_status** | Normal | Both | MDSI | MDSI1 | MDSI3 | -0.1703812 | 0.1019089 | Log odds ratio | 7.332101 |
| **Age** |  |  | MDSI | MDSI1 | MDSI4 | -3.568583 | 0.4143268 | Log odds ratio | 7.282593 |
| **K_status** | Normal | Normal | MDSI | MDSI1 | MDSI4 | -1.350506 | 0.1523519 | Log odds ratio | 7.332101 |
| **K_status** | Normal | Hypo | MDSI | MDSI1 | MDSI4 | -4.032368 | 0.5225778 | Log odds ratio | 7.332101 |
| **K_status** | Normal | Hyper | MDSI | MDSI1 | MDSI4 | -1.152356 | 0.1407853 | Log odds ratio | 7.332101 |
| **K_status** | Normal | Both | MDSI | MDSI1 | MDSI4 | 0.34208680 | 0.2847009 | Log odds ratio | 7.332101 |
| **Age** |  |  | MDSI | MDSI1 | MDSI5 | 0.21237660 | 0.293043 | Log odds ratio | 7.282593 |
| **K_status** | Normal | Normal | MDSI | MDSI1 | MDSI5 | 1.591016 | 0.2398793 | Log odds ratio | 7.332101 |
| **K_status** | Normal | Hypo | MDSI | MDSI1 | MDSI5 | -0.8531771 | 0.3997079 | Log odds ratio | 7.332101 |
| **K_status** | Normal | Hyper | MDSI | MDSI1 | MDSI5 | 0.6021516 | 0.2714108 | Log odds ratio | 7.332101 |
| **K_status** | Normal | Both | MDSI | MDSI1 | MDSI5 | -2.128786 | 0.6596835 | Log odds ratio | 7.332101 |
| **Age** |  |  | MDSI | MDSI1 | MDSI6 | -0.2829441 | 0.3298623 | Log odds ratio | 7.282593 |
| **K_status** | Normal | Normal | MDSI | MDSI1 | MDSI6 | -0.8953012 | 0.141185 | Log odds ratio | 7.332101 |
| **K_status** | Normal | Hypo | MDSI | MDSI1 | MDSI6 | -1.727015 | 0.195295 | Log odds ratio | 7.332101 |
| **K_status** | Normal | Hyper | MDSI | MDSI1 | MDSI6 | -2.464894 | 0.2692077 | Log odds ratio | 7.332101 |
| **K_status** | Normal | Both | MDSI | MDSI1 | MDSI6 | -3.107774 | 0.3648053 | Log odds ratio | 7.332101 |
| **Age** |  |  | MDSI | MDSI1 | MDSI7 | -3.080876 | 0.3627015 | Log odds ratio | 7.282593 |
| **K_status** | Normal | Normal | MDSI | MDSI1 | MDSI7 | -5.524363 | 1.037914 | Log odds ratio | 7.332101 |
| **K_status** | Normal | Hypo | MDSI | MDSI1 | MDSI7 | -1.820408 | 0.20399 | Log odds ratio | 7.332101 |
| **K_status** | Normal | Hyper | MDSI | MDSI1 | MDSI7 | -0.6611451 | 0.6327682 | Log odds ratio | 7.332101 |
| **K_status** | Normal | Both | MDSI | MDSI1 | MDSI7 | -2.035755 | 1.072368 | Log odds ratio | 7.332101 |
| **Age** |  |  | MDSI | MDSI1 | MDSI8 | -0.9069175 | 0.6927384 | Log odds ratio | 7.282593 |
| **K_status** | Normal | Normal | MDSI | MDSI1 | MDSI8 | -1.983614 | 1.071397 | Log odds ratio | 7.332101 |
| **K_status** | Normal | Hypo | MDSI | MDSI1 | MDSI8 | -1.548893 | 0.8223461 | Log odds ratio | 7.332101 |
| **K_status** | Normal | Hyper | MDSI | MDSI1 | MDSI8 | -1.987816 | 1.07584 | Log odds ratio | 7.332101 |
| **K_status** | Normal | Both | MDSI | MDSI1 | MDSI8 | -2.079464 | 1.074774 | Log odds ratio | 7.332101 |
| **ICDSC_max** | 0-2 | 0-2 | Analgesics | 0 | 1 | -0.568793 | 0.1095824 | Log odds ratio<br>(Analgesics=1) | 6.148949 |
| **ICDSC_max** | 0-2 | 3 | Analgesics | 0 | 1 | -0.1836483 | 0.1779556 | Log odds ratio | 6.148949 |
| **ICDSC_max** | 0-2 | ≥4 | Analgesics | 0 | 1 | -0.08758934 | 0.1185837 | Log odds ratio | 6.148949 |
| **SAPS** |  |  | Analgesics | 0 | 1 | -0.5040669 | 0.06007353 | Log odds ratio<br>(Analgesics=1) | 6.148985 |
| **Anesthetics** |  |  | Analgesics | 0 | 1 | 0.7316307 | 0.1187853 | Log odds ratio | 6.156197 |
| **Ventilation** |  |  | Anesthetics | 0 | 1 | 2.289818 | 0.1349553 | Log odds ratio<br>(Anesthetics=1) | 2.807269 |
| **ICD_Delirium** | 0 | 1 | Anesthetics | 0 | 1 | 0.6858772 | 0.1380828 | Log odds ratio | 2.645437 |
| **ICDSC_max** | 0-2 | 3 | Antipsychotics | 0 | 1 | 0.7739542 | 0.2570282 | Log odds ratio | 5.479331 |
| **ICDSC_max** | 0-2 | ≥4 | Antipsychotics | 0 | 1 | 2.177477 | 0.1664075 | Log odds ratio | 5.479331 |
| **MDSI1** | MDSI1 | MDSI1 | Antipsychotics | 0 | 1 | -1.41343 | 0.1942583 | Log odds ratio | 5.414411 |
| **MDSI2** | MDSI1 | MDSI2 | Antipsychotics | 0 | 1 | -1.311554 | 0.2262937 | Log odds ratio | 5.414411 |
| **MDSI3** | MDSI1 | MDSI3 | Antipsychotics | 0 | 1 | -2.477286 | 0.2951096 | Log odds ratio | 5.414411 |
| **MDSI4** | MDSI1 | MDSI4 | Antipsychotics | 0 | 1 | -3.147311 | 0.2476621 | Log odds ratio | 5.414411 |
| **MDSI5** | MDSI1 | MDSI5 | Antipsychotics | 0 | 1 | -3.345778 | 0.7712487 | Log odds ratio | 5.414411 |
| **MDSI6** | MDSI1 | MDSI6 | Antipsychotics | 0 | 1 | -1.58953 | 0.2767546 | Log odds ratio | 5.414411 |
| **MDSI7** | MDSI1 | MDSI7 | Antipsychotics | 0 | 1 | -1.950324 | 0.7082438 | Log odds ratio | 5.414411 |
| **MDSI8** | MDSI1 | MDSI8 | Antipsychotics | 0 | 1 | -1.741065 | 0.274687 | Log odds ratio | 5.414411 |
| **Ventilation** | 0 | 1 | Antipsychotics | 0 | 1 | -0.7613606 | 0.1563082 | Log odds ratio | 5.37688 |
| **SAPS** |  |  | Hyperoxia | None | mild | 0.02980558 | 0.06711527 | Log odds ratio | 6.21421 |
| **Anesthetics** | None | mild | Hyperoxia | None | mild | -0.3625062 | 0.08937246 | Log odds ratio | 6.166788 |
| **Ventilation** | 0 | 1 | Hyperoxia | None | mild | -0.7942053 | 0.1160752 | Log odds ratio | 6.145875 |
| **SAPS** |  |  | Hyperoxia | None | moderate | 0.3741548 | 0.1510403 | Log odds ratio | 6.21421 |
| **Anesthetics** | 0 | 1 | Hyperoxia | None | moderate | 1.106195 | 0.2139141 | Log odds ratio | 6.166788 |
| **Ventilation** | 0 | 1 | Hyperoxia | None | moderate | 2.4427842 | 0.4417843 | Log odds ratio | 6.145875 |
| **SAPS** |  |  | Hyperoxia | None | severe | -0.2919364 | 0.1560283 | Log odds ratio | 6.21421 |
| **Anesthetics** | 0 | 1 | Hyperoxia | None | severe | 0.1145972 | 0.2121432 | Log odds ratio | 6.166788 |
| **Ventilation** | 0 | 1 | Hyperoxia | None | severe | 1.385639 | 0.3801052 | Log odds ratio | 6.145875 |
| **Antipsychotics** |  |  | Hypoxia | None | mild | 0.5738619 | 0.155487 | Log odds ratio | 4.948589 |
| **Hyperoxia** | None | mild | Hypoxia | None | mild | 0.8114891 | 0.2229358 | Log odds ratio | 5.008327 |
| **Hyperoxia** | None | mild | Hypoxia | None | mild | 0.9627042 | 0.2415919 | Log odds ratio | 5.008327 |
| **Hyperoxia** | None | moderate | Hypoxia | None | mild | 0.2732042 | 0.1286752 | Log odds ratio | 5.008327 |
| **Hyperoxia** | None | severe | Hypoxia | None | mild | -1.709485 | 0.2329722 | Log odds ratio | 5.008327 |
| **Ventilation** | 0 | 1 | Hypoxia | None | mild | -1.212842 | 0.2091299 | Log odds ratio | 4.959577 |
| **Antipsychotics** | 0 | 1 | Hypoxia | None | moderate | -0.7476133 | 0.1490179 | Log odds ratio | 4.948589 |
| **Hyperoxia** | None | None | Hypoxia | None | moderate | -2.844613 | 0.2834009 | Log odds ratio | 5.008327 |
| **Hyperoxia** | None | mild | Hypoxia | None | moderate | -2.938594 | 0.3287628 | Log odds ratio | 5.008327 |
| **Hyperoxia** | None | moderate | Hypoxia | None | moderate | -0.9559365 | 0.1993795 | Log odds ratio | 5.008327 |
| **Hyperoxia** | None | severe | Hypoxia | None | moderate | -2.550719 | 0.3218385 | Log odds ratio | 5.008327 |
| **Ventilation** | 0 | 1 | Hypoxia | None | moderate | -2.413303 | 0.3663711 | Log odds ratio | 4.959577 |
| **Antipsychotics** | 0 | 1 | Hypoxia | None | severe | -1.311519 | 0.2571045 | Log odds ratio | 4.948589 |
| **Hyperoxia** | None | None | Hypoxia | None | severe | -2.829204 | 0.3925142 | Log odds ratio | 5.008327 |
| **Hyperoxia** | None | None | Hypoxia | None | severe | -1.839283 | 0.3831986 | Log odds ratio | 5.008327 |
| **Hyperoxia** | None | moderate | Hypoxia | None | severe | 0.5288276 | 0.1352931 | Log odds ratio | 5.008327 |
| **Hyperoxia** | None | Severe | Hypoxia | None | severe | 1.121005 | 0.2364262 | Log odds ratio | 5.008327 |
| **Ventilation** | 0 | 1 | Hypoxia |  |  | 0.03860393 | 0.2299334 | Log odds ratio | 4.959577 |
| **Anesthetics** | 0 | 1 | ICD_Thyroid | 0 | 1 | -0.6584648 | 0.2065962 | Log odds ratio | 1.332056 |
| **MDSI1** | MDSI1 | MDSI1 | ICD_Malnutrition | 0 | 1 | -2.847812 | 0.2099574 | Log odds ratio | 2.989774 |
| **MDSI2** | MDSI1 | MDSI2 | ICD_Malnutrition | 0 | 1 | -1.821612 | 0.1876299 | Log odds ratio | 2.989774 |
| **MDSI3** | MDSI1 | MDSI3 | ICD_Malnutrition | 0 | 1 | -1.059772 | 0.1961439 | Log odds ratio | 2.989774 |
| **MDSI4** | MDSI1 | MDSI4 | ICD_Malnutrition | 0 | 1 | -3.374169 | 0.321597 | Log odds ratio | 2.989774 |
| **MDSI5** | MDSI1 | MDSI5 | ICD_Malnutrition | 0 | 1 | -3.135494 | 1.021508 | Log odds ratio | 2.989774 |
| **MDSI6** | MDSI1 | MDSI6 | ICD_Malnutrition | 0 | 1 | -1.995672 | 0.2848461 | Log odds ratio | 2.989774 |
| **MDSI7** | MDSI1 | MDSI7 | ICD_Malnutrition | 0 | 1 | -2.197225 | 1.054093 | Log odds ratio | 2.989774 |
| **MDSI8** | MDSI1 | MDSI8 | ICD_Malnutrition | 0 | 1 | -1.673976 | 0.2568506 | Log odds ratio | 2.989774 |
| **Sex** |  |  | ICD_Diabetes | 0 | 1 | 0.5852454 | 0.151543 | Log odds ratio | 6.014102 |
| **Age** |  |  | ICD_Diabetes | 0 | 1 | 0.5018253 | 0.07760267 | Log odds ratio | 6.05216 |
| **Analgesics** | 0 | 1 | ICD_Diabetes | 0 | 1 | -0.5311409 | 0.1360533 | Log odds ratio | 6.015811 |
| **PCT** | 0 | 1 | Pain | 0 | 1 | 0.694009 | 0.1678918 | Log odds ratio | 2.563237 |
| **Analgesics** | 0 | 1 | Pain | 0 | 1 | 0.611176 | 0.1157652 | Log odds ratio | 2.563268 |
| **MDSI1** | MDSI1 | MDSI1 | Ventilation | 0 | 1 | 0.9745596 | 0.1071354 | Log odds ratio | 3.437062 |
| **MDSI2** | MDSI1 | MDSI2 | Ventilation | 0 | 1 | 1.594324 | 0.1734247 | Log odds ratio | 3.437062 |
| **MDSI3** | MDSI1 | MDSI3 | Ventilation | 0 | 1 | 0.5423243 | 0.1778424 | Log odds ratio | 3.437062 |
| **MDSI4** | MDSI1 | MDSI4 | Ventilation | 0 | 1 | 0.1060596 | 0.1152489 | Log odds ratio | 3.437062 |
| **MDSI5** | MDSI1 | MDSI5 | Ventilation | 0 | 1 | 0.1670541 | 0.4096732 | Log odds ratio | 3.437062 |
| **MDSI6** | MDSI1 | MDSI6 | Ventilation | 0 | 1 | 0.9769154 | 0.2073999 | Log odds ratio | 3.437062 |
| **MDSI7** | MDSI1 | MDSI7 | Ventilation | 0 | 1 | -0.847297 | 0.6900656 | Log odds ratio | 3.437062 |
| **MDSI8** | MDSI1 | MDSI8 | Ventilation | 0 | 1 | 0.3184537 | 0.1896967 | Log odds ratio | 3.437062 |
| **Sex** | Female | Male | ICD_Delirium | 0 | 1 | 0.6251668 | 0.1498306 | Log odds ratio | 6.935387 |
| **Age** |  |  | ICD_Delirium | 0 | 1 | 0.3665982 | 0.0733981 | Log odds ratio | 6.98443 |
| **ICDSC_max0** | 0-2 | 0-2 | ICD_Delirium | 0 | 1 | -2.408728 | 0.1605834 | Log odds ratio | 7.079296 |
| **ICDSC_max3** | 0-2 | 3 | ICD_Delirium | 0 | 1 | -1.655094 | 0.2178786 | Log odds ratio | 7.079296 |
| **ICDSC_max4** | 0-2 | ≥4 | ICD_Delirium | 0 | 1 | -0.1237106 | 0.1449 | Log odds ratio | 7.079296 |
| **Antipsychotics** | 0 | 1 | ICD_Delirium | 0 | 1 | 1.365641 | 0.1658286 | Log odds ratio | 6.97128 |

Delirium Tremens
| Parent | from | to | Child | from | to | Coefficient |
| --- | --- | --- | --- | --- | --- | --- |
| **Sex** | female | male | K_status | normal | hypo | -0.33311 |
| **Age** |  |  | K_status | normal | hypo | 0.676678 |

|  |  |  |  |  |  |  |
| --- | --- | --- | --- | --- | --- | --- |
| **Sex** | female | male | **K_status** | normal | hyper | 0.36363 |
| **Age** |  |  | **K_status** | normal | hyper | -0.37161 |
| **Sex** | female | male | **K_status** | normal | both | 0.249568 |
| **Age** |  |  | **K_status** | normal | both | -0.53899 |
| **Age** |  |  | **alb_status** | normal | hypo | 0.913911 |
| **SAPS** |  |  | **alb_status** | normal | hypo | 1.023615 |
| **Antipsychotics** | 0 | 1 | **ICDSC_max** | 0-2 | 3 | 0.956293 |
| **Antipsychotics** | 0 | 1 | **ICDSC_max** | 0-2 | ≥4 | 1.867205 |
| **Sex** | female | male | **PCT** | 0 | 1 | -0.54889 |
| **Age** |  |  | **PCT** | 0 | 1 | -0.55506 |
| **SAPS** |  |  | **PCT** | 0 | 1 | 0.462619 |
| **Age** |  |  | **ICD_Dementia** | 0 | 1 | 2.346699 |
| **Age** |  |  | **SAPS** |  |  | 0.322703 |
| **K_status** | normal | normal | **SAPS** |  |  | 0.043533 |
| **K_status** | normal | hypo | **SAPS** |  |  | -0.3294 |
| **K_status** | normal | hyper | **SAPS** |  |  | 0.148472 |
| **K_status** | normal | both | **SAPS** |  |  | -0.60185 |
| **Na_status** | normal | normal | **SAPS** |  |  | -0.11366 |
| **Na_status** | normal | hypo | **SAPS** |  |  | -0.2953 |
| **Na_status** | normal | hyper | **SAPS** |  |  | 0.231236 |
| **Na_status** | normal | both | **SAPS** |  |  | -0.56153 |
| **ICD_Malnutrition** | 0 | 1 | **SAPS** |  |  | 0.317396 |
| **K_status** | normal | normal | **MDSI** | MDSI1 | MDSI2 | -3.09147 |
| **K_status** | normal | hypo | **MDSI** | MDSI1 | MDSI2 | -2.00593 |
| **K_status** | normal | hyper | **MDSI** | MDSI1 | MDSI2 | -1.34272 |
| **K_status** | normal | both | **MDSI** | MDSI1 | MDSI2 | -3.77483 |
| **PCT** | 0 | 1 | **MDSI** | MDSI1 | MDSI2 | -2.52889 |
| **Ventilation** | 0 | 1 | **MDSI** | MDSI1 | MDSI2 | -24.3784 |
| **K_status** | normal | normal | **MDSI** | MDSI1 | MDSI3 | -2.92735 |
| **K_status** | normal | hypo | **MDSI** | MDSI1 | MDSI3 | -1.92093 |
| **K_status** | normal | hyper | **MDSI** | MDSI1 | MDSI3 | -0.97769 |
| **K_status** | normal | both | **MDSI** | MDSI1 | MDSI3 | 0.4294 |

|  |  |  |  |  |  |  |
| --- | --- | --- | --- | --- | --- | --- |
| ICDSC_max0 | 0-2 | 0-2 | Analgesics | 0 | 1 | -0.51134 |
| ICDSC_max3 | 0-2 | 3 | Analgesics | 0 | 1 | -0.02077 |
| ICDSC_max4 | 0-2 | ≥4 | Analgesics | 0 | 1 | 0.166777 |
| SAPS |  |  | Analgesics | 0 | 1 | -0.5181 |
| Anesthetics | 0 | 1 | Analgesics | 0 | 1 | 0.725354 |
| Na_status | normal | hypo | Anesthetics | 0 | 1 | -0.55318 |
| Na_status | normal | hyper | Anesthetics | 0 | 1 | 0.750999 |
| Na_status | normal | both | Anesthetics | 0 | 1 | 5.288662 |
| SAPS |  |  | Anesthetics | 0 | 1 | 0.447725 |
| Hyperoxia | none | none | Anesthetics | 0 | 1 | -0.30965 |
| Hyperoxia | none | mild | Anesthetics | 0 | 1 | 0.051976 |
| Hyperoxia | none | moderate | Anesthetics | 0 | 1 | 1.06279 |
| Hyperoxia | none | severe | Anesthetics | 0 | 1 | 3.081941 |
| ICD_DTs | 0 | 1 | Anesthetics | 0 | 1 | 1.814247 |
| SAPS | none | mild | Hyperoxia | none | mild | -0.01063 |
| SAPS | none | moderate | Hyperoxia | none | moderate | -0.27065 |
| SAPS | none | severe | Hyperoxia | none | severe | -0.65462 |
| Hyperoxia | none | none | Hypoxia | none | mild | 0.627542 |
| Hyperoxia | none | mild | Hypoxia | none | mild | -0.77584 |
| Hyperoxia | none | moderate | Hypoxia | none | mild | -1.05209 |
| Hyperoxia | none | severe | Hypoxia | none | mild | -0.35437 |
| Hyperoxia | none | none | Hypoxia | none | moderate | -2.11224 |
| Hyperoxia | none | mild | Hypoxia | none | moderate | -3.434 |
| Hyperoxia | none | moderate | Hypoxia | none | moderate | -0.69315 |
| Hyperoxia | none | severe | Hypoxia | none | moderate | -1.82162 |
| Hyperoxia | none | none | Hypoxia | none | severe | -2.61009 |
| Hyperoxia | none | mild | Hypoxia | none | severe | -0.81094 |
| Hyperoxia | none | moderate | Hypoxia | none | severe | -1.72724 |
| Hyperoxia | none | severe | Hypoxia | none | severe | -1.86077 |
| Sex |  |  | ICD_Diabetes | 0 | 1 | 0.618342 |
| Age |  |  | ICD_Diabetes | 0 | 1 | 0.588398 |
| Analgesics | 0 | 1 | ICD_Diabetes | 0 | 1 | -0.51969 |

|  |  |  |  |  |  |  |
| --- | --- | --- | --- | --- | --- | --- |
| **PCT** | 0 | 1 | Pain | 0 | 1 | 0.673716 |
| **Analgesics** | 0 | 1 | Pain | 0 | 1 | 0.656674 |
| **SAPS** |  |  | Ventilation | 0 | 1 | 0.688998 |
| **Anesthetics** | 0 | 1 | Ventilation | 0 | 1 | 3.048348 |
| **Hypoxia** | none | none | Ventilation | 0 | 1 | -0.99826 |
| **Hypoxia** | none | mild | Ventilation | 0 | 1 | -0.54278 |
| **Hypoxia** | none | moderate | Ventilation | 0 | 1 | 1.197175 |
| **Hypoxia** | none | severe | Ventilation | 0 | 1 | -0.61294 |
| **ICD_DTs** | 0 | 1 | Ventilation | 0 | 1 | -1.5069 |
| **ICDSC** | 0-2 | 0-2 | ICD_DTs | 0 | 1 | -4.76899 |
| **ICDSC** | 0-2 | 3 | ICD_DTs | 0 | 1 | -4.7185 |
| **ICDSC** | 0-2 | ≥4 | ICD_DTs | 0 | 1 | -2.00271 |

Children
| Parameter from | State 0 | State 1 | To | State 0 | State 1 | Coefficient | Standard Error | Interpretation | Arc-strength |
| --- | --- | --- | --- | --- | --- | --- | --- | --- | --- |
| **Amm** | Normal | Low | ICD_HE | 0 | 1 | -3.479748 | NA | Log odds ratio | 3.52 |
| **Amm** | normal | High | ICD_HE | 0 | 1 | 0.748449 | 0.3186329 | Log odds ratio | 3.52 |
| **ICDSC_max** | 0-2 | 0-2 | ICD_HE | 0 | 1 | -1.827529 | 0.3003714 | Log odds ratio | 3.47 |
| **ICDSC_max** | 0-2 | 3 | ICD_HE | 0 | 1 | -1.214438 | 0.5373295 | Log odds ratio | 3.47 |
| **ICDSC_max** | 0-2 | ≥4 | ICD_HE | 0 | 1 | -0.7100362 | 0.2929244 | Log odds ratio | 3.47 |
| **Antipsychotics** | 0 | 1 | ICDSC_max | 0-2 | 3 | -7.10146 | 28.20183 | Log odds ratio | 1.78 |
| **Antipsychotics** | 0 | 1 | ICDSC_max | 0-2 | ≥4 | 1.227331 | 0.4617435 | Log odds ratio | 1.78 |
| **SAPS** |  |  | PCT | 0 | 1 | 1.090322 | 0.3430428 | Log odds ratio | 3.13 |
| **ICD_HE** | 0 | 1 | SAPS |  |  | 0.549457 | NA | Correlation | 7.96 |
| **Age** |  |  | SAPS |  |  | 0.2371777 | NA | Correlation | 8.73 |
| **K_status** | Normal | Both | SAPS |  |  | 0.002313853 | NA | Correlation | 8.25 |
| **K_status** | Normal | Hyper | SAPS |  |  | -0.08863074 | NA | Correlation | 8.25 |
| **K_status** | Normal | Hypo | SAPS |  |  | -0.5675481 | NA | Correlation | 8.25 |
| **K_status** | Normal | Normal | SAPS |  |  | -0.1908517 | NA | Correlation | 8.25 |
| **Na_status** | Normal | Both | SAPS |  |  | -0.2049992 | NA | Correlation | 8.18 |
| **Na_status** | Normal | Hyper | SAPS |  |  | -0.09668247 | NA | Correlation | 8.18 |
| **Na_status** | Normal | Hypo | SAPS |  |  | -0.3439229 | NA | Correlation | 8.18 |
| **Na_status** | Normal | Normal | SAPS |  |  | -0.1991121 | NA | Correlation | 8.18 |
| **Ventilation** | 0 | 1 | SAPS | 0 | 1 | 0.4481908 | NA | Correlation | 7.97 |
| **SAPS** |  |  | Analgesics | 0 | 1 | -0.3712373 | 0.1347155 | Log odds ratio | 4.68 |
| **Anesthetics** | 0 | 1 | Analgesics | 0 | 1 | 0.9320333 | 0.2814237 | Log odds ratio | 4.68 |
| **Ventilation** | 0 | 1 | Anesthetics | 0 | 1 | 2.399075 | 0.3220783 | Log odds ratio | 1.8 |

